# Leveraging Limited Testing Data for Early Detection of Emerging Infectious Disease Outbreaks

**DOI:** 10.64898/2025.12.22.25342841

**Authors:** Alexander John Zapf, Marc Lipsitch

## Abstract

Early during emerging infectious disease outbreaks, case-based surveillance is constrained by limited test availability, diagnostic delays, and low clinical suspicion. Novel pathogens that mimic symptoms of established conditions may generate detectable outbreak signals in routine testing data, as infected individuals seek testing for known conditions and test negative. We developed analytic and simulation frameworks using Poisson and negative binomial models to evaluate whether total and negative testing volumes for a clinically similar (“mimicking”) condition can provide timely outbreak warning. We systematically assessed detection performance across variations in baseline test counts, epidemic growth rates, testing fractions among cases, detection threshold stringency, and overdispersion. Detection thresholds based on negative tests consistently outperformed total test thresholds, achieving earlier detection with approximately one-third fewer cumulative cases while maintaining comparable false positive rates. However, reliable detection required stringent conditions: high testing fractions among epidemic cases, low baseline test volumes, and low overdispersion. When testing fractions fell below 5%, epidemic size estimates provided little practical information; above 30%, precision markedly improved. These findings support prioritizing access to disaggregated test result data but caution that this detection approach is best positioned as a resource-efficient complement within integrated surveillance portfolios rather than a standalone early warning system.

## 1. Introduction

Early warning for emerging infectious disease outbreaks requires public health surveillance systems that can recognize a novel threat before case-based reporting is established. Recent emergencies — the COVID-19 pandemic, the 2022 mpox outbreaks, and the 2009 H1N1 pandemic — have highlighted that when a novel pathogen clinically resembles a routinely tested condition, conventional surveillance based on confirmed cases can be blind for weeks: testing is unavailable or very limited, diagnostic delays accumulate, and clinical suspicion is low.¹ Yet patients infected with the novel pathogen are already entering the healthcare system and being tested for the mimicking disease.

Against the backdrop of growing bioterrorism and health security threats, surveillance using *syndromic* data, based on a clinical encounter without confirmation of the cause of illness, has received increasing attention and development since the early 2000s.^1–5^ More recent developments use non-traditional data streams, including wastewater surveillance, mobility patterns, and automated aberration detection algorithms scanning information from various large-scale databases.^2,6–13^ Many of these data streams, including diagnostic testing volumes, are generated and collected at an unprecedented scale amidst the expansion of health management and information systems globally. Nonetheless, assessments of the usefulness of such data for disease surveillance purposes remain limited.^2,14,15^

Leveraging signals of health threats from alternative data streams has the potential to improve the timeliness of outbreak detection while keeping false alarms at manageable levels.^1,2,15,16^ For instance, changes in the total volumes of tests for similar disease syndromes, such as influenza tests at the onset of the COVID-19 pandemic^17,18^ or syphilis tests during mpox outbreaks,^19,20^ may be leveraged as indicators that signal emerging infections.

Identifying the conditions under which test volumes can provide reliable and timely warning of new infectious disease outbreaks is a precursor to designing and deploying such systems in settings where other information is limited, potentially in combination with other data streams to promote the integration of diverse data into a surveillance framework akin to a “mosaic.”^21,22^

This research seeks to address this gap by systematically evaluating the performance of using total and negative testing volume data for a mimicking condition as an early indicator for an emerging infectious disease with little or no available surveillance data. The primary goal is to identify the key epidemiologic parameters and conditions under which this surveillance approach can enable reliable outbreak detection, thereby contributing to enhanced public health intelligence and preparedness infrastructures.

## 2. Methods

### 2.1 Heuristic Overview

Emerging infectious disease outbreaks driven by novel pathogens may mimic symptoms of well-monitored conditions for which testing is available. This similarity creates opportunities for early outbreak detection: patients infected with the novel “mimic” pathogen may undergo testing for the known condition (and typically test negative), generating an outbreak signal within routine testing data. Leveraging such signals for early disease detection therefore hinges on isolating the contribution of emerging outbreaks from ordinary variation in routine test counts.

This phenomenon has been observed across diverse diseases and epidemic contexts (Table 1). The 2009 A(H1N1) pandemic influenza strain was first identified in specimens from patients with influenza-like illness (ILI) that were positive for influenza A but could not be subtyped as circulating seasonal influenza.^23–25^ The first community transmission of SARS-CoV-2 in the US was identified by testing specimens from ILI surveillance, and retrospective analyses have shown that outliers in influenza-negative ILI predated reported COVID-19 surges by multiple weeks across 16 countries during the early COVID-19 pandemic.^26–28^ During the 2022-2023 mpox outbreak, genital lesions closely resembling primary syphilis led to widespread STI testing among monkeypox-infected individuals, many of whom tested negative for syphilis.^19,20,29,30^ Other documented mimicking scenarios include Nipah virus infections initially misdiagnosed as Japanese encephalitis,^31–33^ Zika virus outbreaks identified via dengue surveillance when suspected dengue cases tested negative,^34–36^ and the use of syndromic surveillance systems to detect bioterrorism agents^4,5,37^ (cf. Supplement A).

**Table 1:** Novel Infection – Mimicking Condition Pairs with Key Testing Data Streams and Mechanisms for Detection (cf. Supplement A)

| **Novel infection** | **Mimicking (known) condition** | **Testing data stream** | **Key detection mechanism** | **References** |
| --- | --- | --- | --- | --- |
| **Mpox** | **STI (syphilis, HSV)** | STI testing volumes | Negative syphilis/ HSV tests in genital lesion workup | 19,20,29,30,47,63–64 |
| **Pandemic influenza** | **Seasonal influenza** | ILI surveillance + influenza subtyping | Unsubtypable influenza A specimens (especially outside of influenza season) | 23–25,65–68 |
| **COVID-19** | **Influenza/ ILI** | ILI surveillance + virologic testing | Influenza-negative surges in ILI | 17,18,26–28,45,46 |
| **Neurotropic pathogens (EV-D68, WNV)** | **Polio (AFP surveillance)** | AFP surveillance/ case reports + poliovirus testing | Polio-negative AFP cases | 69–74 |
| **Smallpox / novel poxviruses** | **Chickenpox (varicella)/ mpox** | Varicella surveillance/ syndromic surveillance for rash | VZV-negative vesicular rash | 29,62,75–80 |
| **Bioterrorism agents (anthrax, plague, tularemia)** | **Syndromes (ILI, pneumonia)** | Syndromic surveillance | Culture/ test-negative severe illness | 4,5,81–84 |
| **Nipah (Hendra) virus** | **Japanese encephalitis** | Encephalitis surveillance | JE-negative encephalitis cases | 31–33,85–87 |
| **Novel respiratory pathogens of pandemic potential** | **COVID-19/ influenza/ RSV** | Multiplex respiratory panels | Pan-negative respiratory panel results | 21,22,45,50,58,88,89 |
| **Zika virus** | **Dengue fever** | Dengue surveillance | DENV-negative febrile rash illness | 34–36,90,91 |
| **Ebola/ VHF** | **Malaria/ febrile illness** | Malaria RDT/ febrile illness surveillance | Malaria-negative severe febrile illness | 83,92–95 |

**\*Abbreviations:** STI: Sexually transmitted infection; HSV: Herpes simplex virus; ILI: influenza-like illness; AFP: Acute flaccid paralysis; EV-D68: Enterovirus D68; WNV: West Nile virus; VZV: Varicella zoster virus; JE: Japanese encephalitis; RSV: respiratory syncytial virus; DENV: dengue virus; VHF: viral hemorrhagic fever; RDT: Rapid diagnostic test

We formalize this detection problem as identifying when observed test counts exceed a statistically derived baseline threshold representing expected (“usual”) testing variability. Focusing detection on negative tests provides a more informative signal as a novel disease (e.g., mpox) disproportionately contributes to negative test counts compared to total tests (e.g., for syphilis). Given a fraction (*f*) of tests that are routinely negative, the detection margin for negative tests scales approximately as √𝑓 relative to total tests, while the outbreak signal is nearly fully preserved in the negative stream. This implies that for a fixed threshold for calling a deviation κ (which determines the false alarm rate) and fixed epidemic growth rate of the novel pathogen, focusing on negative tests will detect the new pathogen earlier and at lower case numbers. This difference will be greatest if most tests for the existing pathogen are positive.

### 2.2 Detection Framework

Our approach models routine test counts for a known condition as the sum of independent positive and negative testing components, each with a respective mean (μ) and variance (σ^2^ = μ x φ), where the variance-to-mean ratio (VMR—φ) captures overdispersion commonly observed in surveillance data.^38–40^ When variance equals the mean (φ = 1), the model reduces to a Poisson process, for which closed-form solutions are available (Supplement B). Under overdispersion (φ > 1), we model each baseline component using a negative binomial distribution with mean μ and VMR φ.

After epidemic emergence at time τ, incident cases of the novel pathogen arise following exponential growth with rate ρ. A fraction π of epidemic cases undergoes testing for the known condition, contributing additional tests – assumed to be negative – to the observed count. This augments the total mean test count over time as:

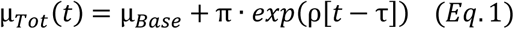

where the expected total baseline test count, 𝜇_𝐵𝑎𝑠𝑒_ = 𝜇_𝑃𝑜𝑠_ + 𝜇_𝑁𝑒𝑔_, is assumed to be stable.

### 2.3 Detection Threshold and Outbreak Detection

An outbreak is detected when the observed test count exceeds a baseline threshold defined as:

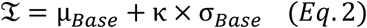

Where σ_Base_ is the standard deviation of total baseline tests and κ is a sensitivity parameter balancing detection timeliness with false positives. Assuming that positive and negative baseline tests arise from independent processes, the total baseline variance is the sum of component variances:

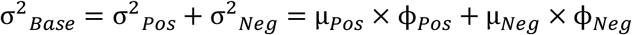

#### 2.3.1 Negative Test Threshold

In secondary analyses, we restrict the detection threshold to negative test counts alone, assuming novel disease cases exclusively contribute negative tests:

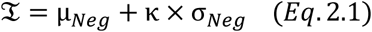

Assuming that additional tests due to novel disease cases are negative, the epidemic signal is preserved but compared to a smaller baseline. This enhances the signal-to-noise ratio and detection can occur earlier, particularly when the proportion of negative tests is small (i.e., when tests for the known condition are frequently positive).

### 2.4 Detection Probability & Time-to-Detection Estimates

Because the sum of negative binomial random variables does not in general follow a negative binomial distribution, we derive the composite distribution of total test counts at each time by sequentially convolving the probability mass functions of the baseline and epidemic test components (cf. Supplement C). From these composite distributions, we compute three key metrics: i) the per-period detection probability, ℙ_𝐷𝑒𝑡_(𝑡), which is the probability that the observed test count at time *t* exceeds the detection threshold; ii) the cumulative detection probability, 𝐹_𝐷𝑒𝑡_(𝑡), representing the probability of detection by time *t* while accounting for all preceding opportunities; and iii) the first-detection distribution, ℙ_𝐷𝑒𝑡1_ (𝑡), which is the probability that detection occurs at exactly time *t*. Based on these metrics, the median detection time (t_50_) is defined as the earliest *t* at which 𝐹_𝐷𝑒𝑡_(𝑡) exceeds 0.5. The expected detection time and expected cumulative cases at detection are computed as weighted sums over the first-detection distribution, ℙ_𝐷𝑒𝑡1_ (𝑡) (formal definitions are provided in Supplement equations B3-B8). Closed-form Poisson expressions and negative binomial convolutions for these metrics are given in Supplement B and C.

### 2.5 Cumulative Epidemic Cases at Detection

Cumulative epidemic cases up to the median detection time (*t_50_*) are estimated by summing the incident case counts from the exponential growth model up to *t_50_*. Incidence (the expected count of new epidemic cases at time *u*) is computed as:

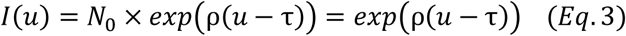

where N_0_ denotes the initial number of novel disease cases (assumed to be N_0_=1) and ρ represents the growth rate of the emerging epidemic.

Cumulative epidemic cases from emergence (τ) to time *t* follow from this exponential growth model:

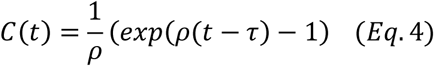

Since the first detection time (𝑡_𝑑_) is a random variable, the *expected cumulative case count* at detection is derived as the weighted average of *C(t)* over the first-detection distribution in post-epidemic periods *t ≥ τ* (cf. Supplement B5).

### 2.6 Epidemic Size Estimation from Routine Data

We developed methods to infer the underlying epidemic size (*N_Epi_*) from routinely observed tests (*X_Obs_*), given a known baseline mean (*μ_Base_*) and testing fraction (π), i.e., the per-case probability of testing for the known condition. The maximum likelihood estimate for the underlying epidemic size (*N̂_Epi_*) is:

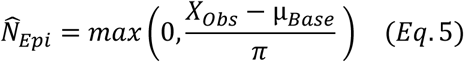

This estimator is independent of baseline variance assumptions (identical under Poisson and NB models). To quantify uncertainty while accounting for overdispersion, we employed a simulation-based likelihood approach. For each hypothetical epidemic size across an adaptive grid, we generated 10,000 simulated test counts by combining negative binomial baseline draws with Poisson epidemic contributions and subsequently estimated the likelihood profile using kernel density estimation. Confidence intervals were derived by test inversion at the α = 0.05 level and validated using parametric bootstraps and likelihood ratio-compatible sets (see Supplement D for details).

### 2.7 Exploration of Parameter Ranges

Key parameters—mean baseline test counts (μ_Pos_ and μ_Neg_), variance-to-mean ratios (φ_Pos_, φ_Neg_), epidemic growth rate (ρ), testing fraction (π), and detection threshold multiplier (κ)—were systematically varied (Table 2), both individually and across multidimensional parameter grids. We corroborated the analytic results through stochastic simulations (cf. Supplement F).

**Table 2:** Parameter Definitions and Assessed Parameter Values.

| Parameter | Definition | Baseline (reference) value | Value range | Extended value range |
| --- | --- | --- | --- | --- |
| $\mu_{\text{Positive}}$ | Mean count of *positive* baseline tests per time unit <sup>a</sup> | **100** | {30, 100, 300, 1000} | {100, 500, 1000, 2000, 5000} |
| $\phi_{\text{Positive}}$ | Variance-to-mean ratio of *positive* baseline tests <sup>b</sup> | **1.5<sup>c</sup>** | {1.5, 3, 5}<sup>c</sup> | {1.5, 3, 5, 10}<sup>c</sup> |
| $\mu_{\text{Negative}}$ | Mean count of *negative* baseline tests per t | **100** | {30, 100, 300, 1000} | {100, 500, 1000, 2000, 5000} |
| $\phi_{\text{Negative}}$ | Variance-to-mean ratio of *negative* baseline tests <sup>b</sup> | **1.5<sup>c</sup>** | {1.5, 3, 5}<sup>c</sup> | {1.5, 3, 5, 10}<sup>c</sup> |
| $\pi$ | Fraction of epidemic cases getting tested for the known condition (all epidemic cases are assumed to test negative) | **0.10** | {0.10, 0.15, 0.20} | {0.05, 0.10, 0.50, 0.70} |
| $\rho$ | Epidemic growth rate of the unknown disease per time unit | **0.20** | {0.15, 0.20, 0.225} | {0.05, 0.10, 0.15, 0.25, 0.35} |
| $\kappa$ | Detection threshold multiplier | **3** | {2, 3, 4} | {3, 5, 7, 10} |

<sup>a</sup> For Poisson models (Supplement B), the mean (expected) count of baseline tests is defined as $\lambda$ .
<sup>b</sup> For Poisson-based approaches, the variance-to-mean ratio parameters collapse to $\phi = 1$ .
<sup>c</sup> The assessed variance-to-mean ratio (overdispersion) range was selected to represent plausible surveillance conditions and corroborated via empirical analyses of publicly available respiratory virus testing data at operational scales where this approach would be deployed (cf. Supplement E).

To quantify uncertainty under various outbreak scenarios, we derived predictive intervals (95% PI) for expected detection times and cumulative case counts from the first-detection probability distributions (ℙ_𝐷𝑒𝑡1_ (𝑡)) as 2.5/50/97.5% quantiles, normalized over t ≥ τ to exclude pre-emergence false alarms.

For each scenario we compared the negative-test threshold to the total-test threshold on three surveillance system attributes: timeliness (expected time and cumulative cases before first alert), false-alert rate (1–specificity), and the precision of the epidemic-size estimate at the time of alert.

Analyses were performed using R 4.5.1^41^.

## 3. Results

### 3.1 Detection Time Distributions

Across scenarios, the expected time from epidemic emergence to first alert (the system’s lead time) and the accompanying median detection time differed by 1–2 reporting periods — days or weeks depending on the cadence of the underlying surveillance system — with higher overdispersion widening the distribution of lead times and extending the right tail of late alerts (Tables 3 & 4). Analytic and simulation results were congruent: median detection times were identical and expected (mean) detection times aligned almost completely (Tables S1 & S2).

Detection based on negative test counts consistently yielded earlier detections than thresholds based on aggregate (total) test counts (Figure 1) and reduced outbreak size at detection by approximately 33%, while maintaining comparable false positive rates (8.5%-10.7 % vs. 10.7%-12.9%, Tables S1 & S2) under reference parameter values (e.g., κ=3; φ=1.5). This is expected, since for any given number of (negative) tests due to the novel infection, the proportional increase in tests will be greater compared to a reference of only negative tests than to all baseline tests. The departure driven by the underlying epidemic is thus more likely to generate a signal above the threshold based on lower negative test counts.

**Figure 1:**
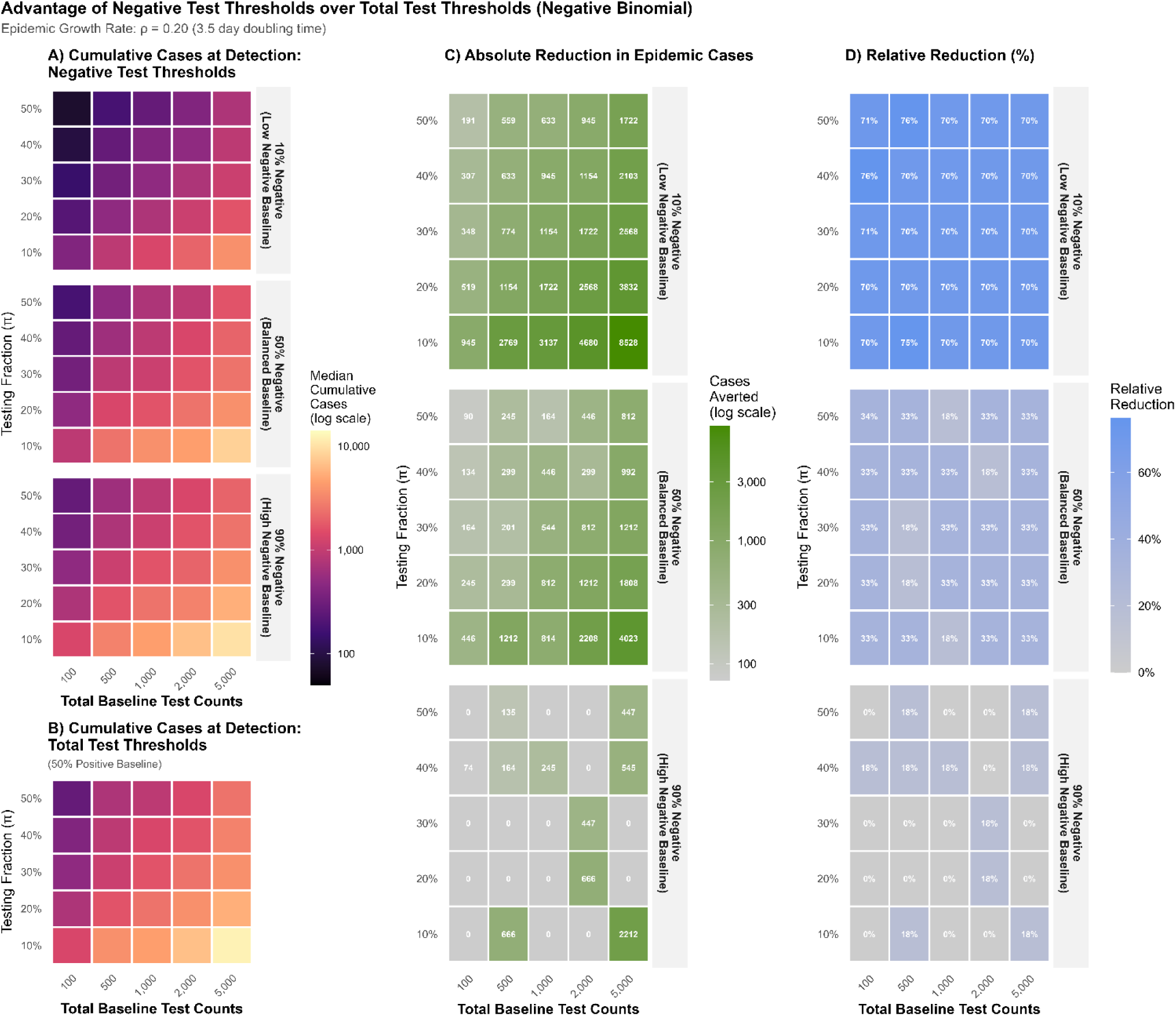
Comparative Detection Performance of Negative versus Total Test Thresholds Under Negative Binomial Assumptions. *Legend*: Heatmaps showing epidemic detection performance across surveillance parameters for two threshold strategies. **Panel A** (Negative Test Thresholds): Detection based on negative test counts for the routine condition. Color gradients indicate median cumulative epidemic cases at detection (log scale: dark purple = early detection with fewer cases; yellow = late detection with more cases). Rows represent increasing baseline proportions of negative tests, demonstrating performance across different baseline testing patterns. The x-axis shows total baseline test counts; the y-axis shows the testing fraction (π), representing the proportion of individuals with the epidemic condition who also receive the routine diagnostic test. **Panel B** (Total Test Thresholds): Detection based on total test counts. **Panel C** (Absolute Case Reduction): Absolute difference in median cumulative cases at detection between strategies. Darker colors (green) indicate larger benefit. **Panel D** (Relative Case Reduction): Percentage reduction in median cumulative cases at detection when using negative test thresholds compared to total test thresholds, calculated as (cases averted / cases at detection using total tests) × 100%. Positive percentages indicate proportional improvement from the negative test strategy. Rows show results stratified by baseline negative test proportions. All panels display results for epidemic growth rate ρ = 0.20 (3.5-day doubling time). X-axes show total baseline test count; Y-axes show testing probability (π). See supplementary *Figure S1* for analogous estimates based on Poisson models and *Figure S2 – S5* for detailed heatmaps displaying estimates of median cumulative epidemic cases at detection.

### 3.2 Influence of Key Parameters and Epidemiologic Conditions

Timeliness and specificity of detections were sensitive to the epidemic growth rate (ρ), the fraction of epidemic cases testing for the known condition (π), and the detection threshold multiplier (κ). Accelerated epidemic growth (increasing ρ from 0.15 to 0.225) reduced detection times by ∼30% (from ∼37 to ∼26 reporting periods) and reduced expected cumulative case counts at detection by ∼20% (from 2410 to 1912 cases), while narrowing the upper tail of the prediction interval (from a 95%PI upper bound of 44 to 30 reporting periods; Tables 3 & 4). Elevated testing fractions produced the largest improvements in detection: doubling π from 0.10 to 0.20 approximately halved expected cases at detection (2047 → 1029 cases; Table 4), reflecting that π directly determines how much of the epidemic signal is captured in observable test counts. Raising the detection threshold multiplier reduced timeliness for greater specificity: low thresholds (κ=2) achieved early detections but with 70%-77% false positives and skewed detection time distributions (wide 95%PI with early tails), whereas high thresholds (κ=4) delayed detection (by 13-14 reporting periods vs. κ=2) while reducing false positives to 0.3%-0.9% (Tables S1 & S2).

Greater variability in test counts – parameterized by VMR (φ) – delayed detection and widened prediction intervals by jointly raising detection thresholds and the spread around expected signals, relative to an idealized Poisson reference (φ=1). False positive rates increased modestly under overdispersion but remained below ∼13% for κ≥3. At κ=2, results under Poisson and negative binomial assumptions converged, suggesting that overdispersion effects diminish where false-positive rates are high. At higher thresholds they diverge as overdispersion inflates the baseline test count distribution that sets the detection threshold. In conjunction with higher baseline test counts (μ_Pos_, μ_Neg_), increased overdispersion (φ) thus further amplified detection delays (Tables 3, 4, S1, S2).

**Table 3:**
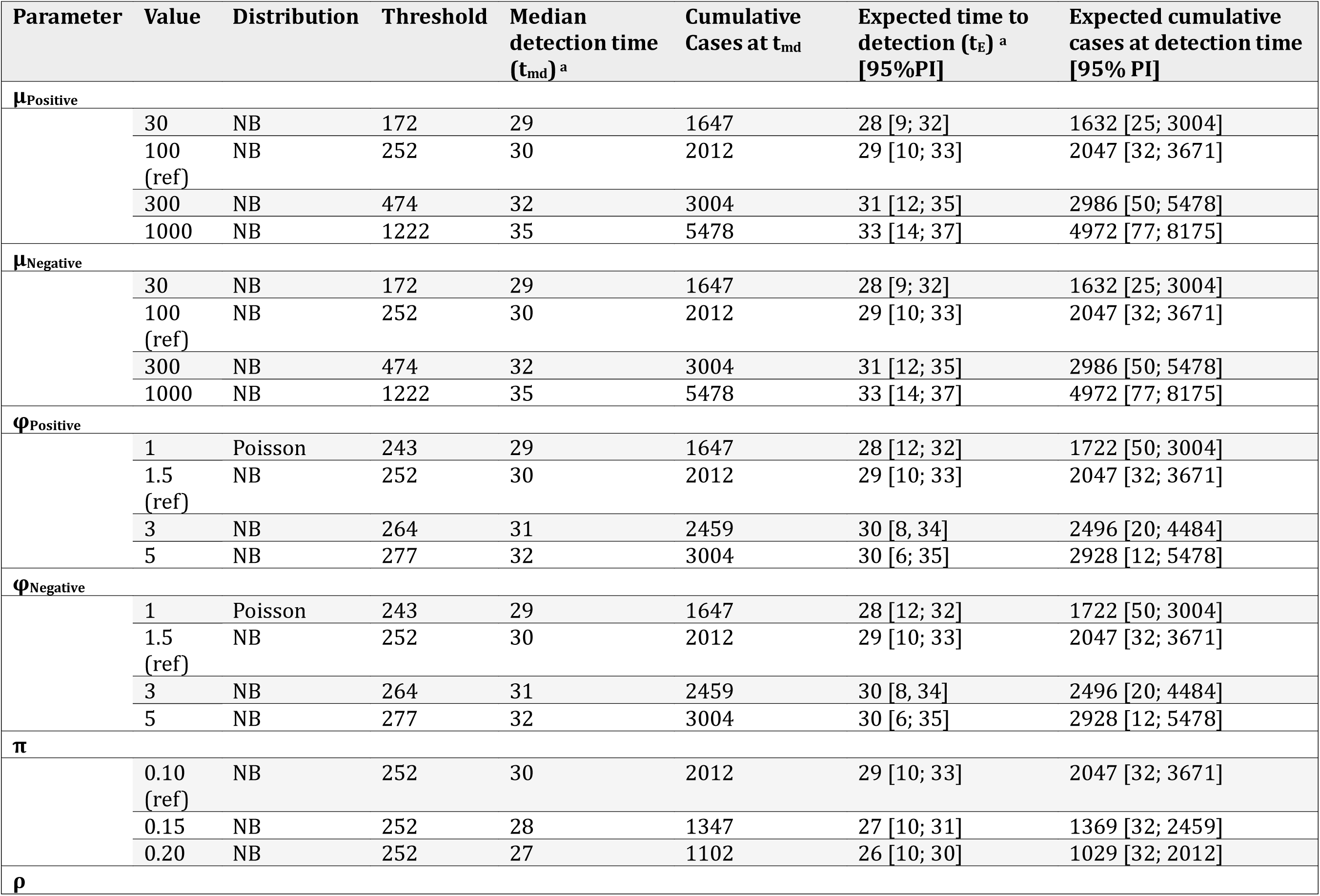
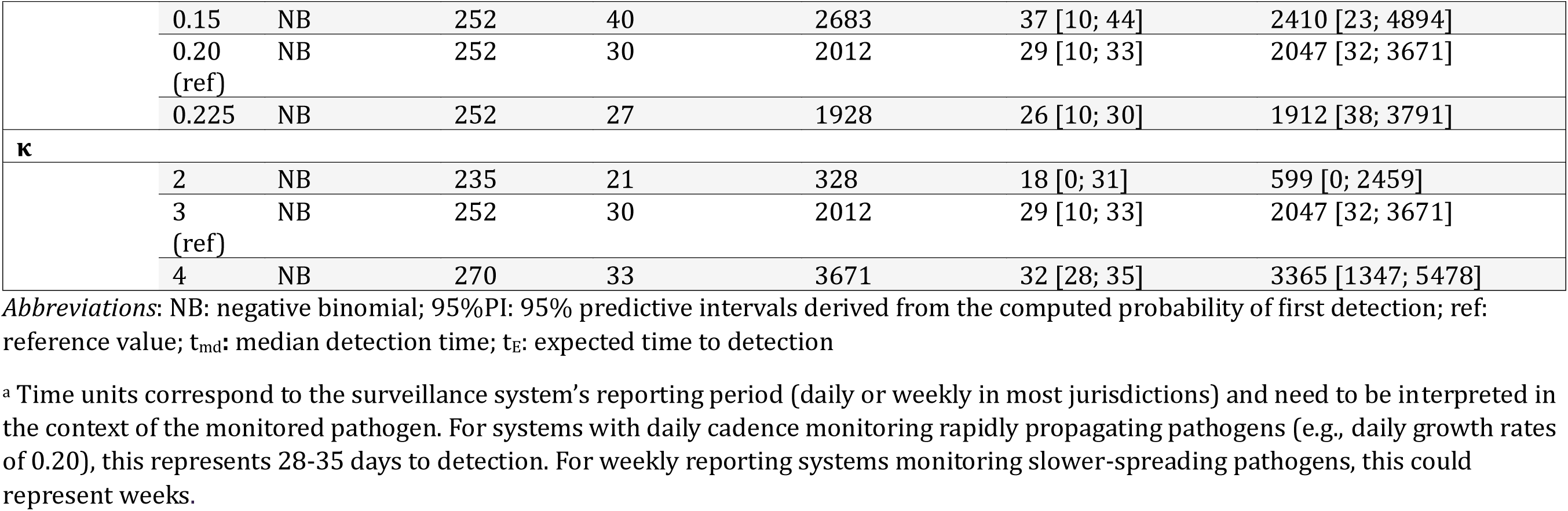
Analytic Estimates of Outbreak Detection Performance Using Total Test Volume Thresholds.

**Table 4:**
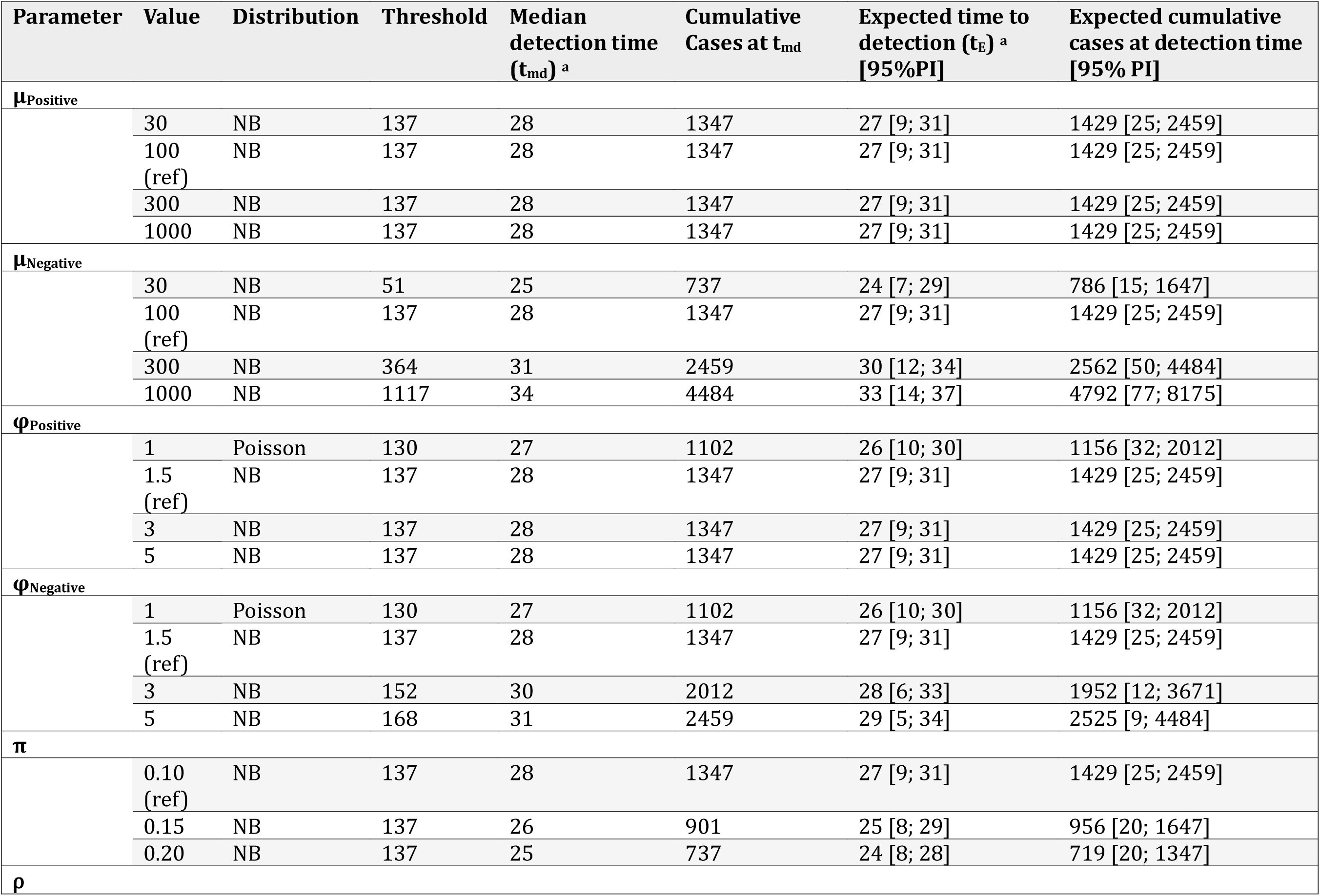
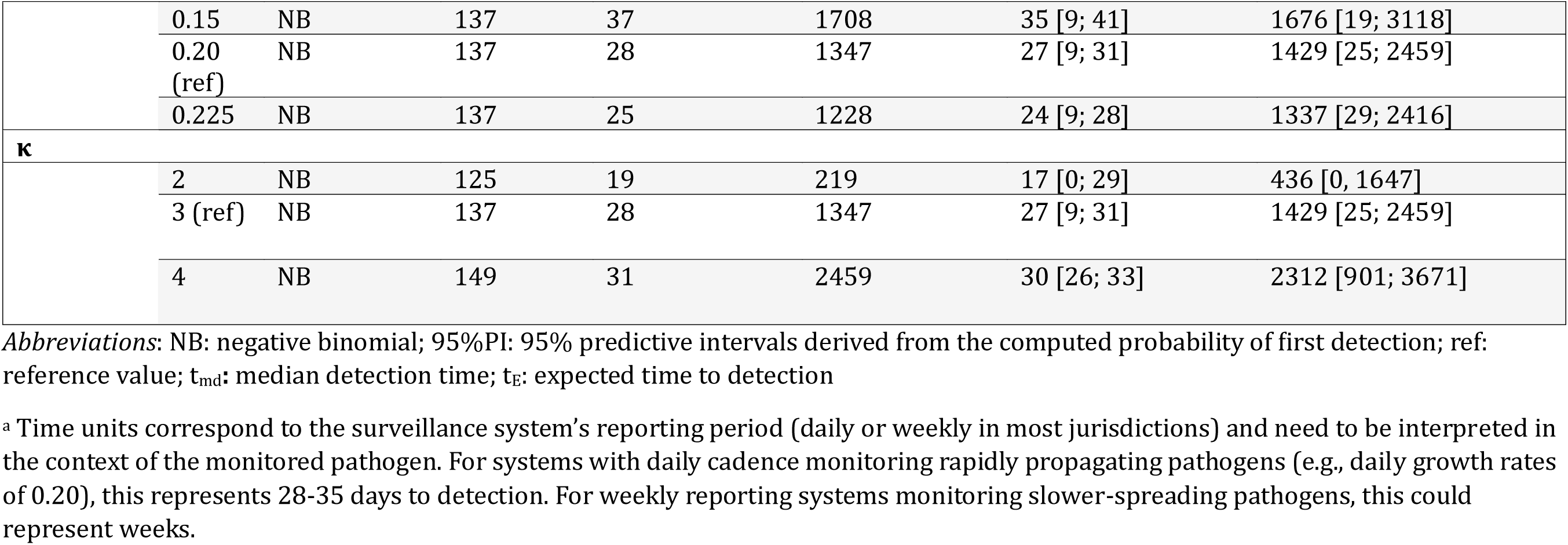
Analytic Estimates of Outbreak Detection Performance Using Negative Test Volume Thresholds.

### 3.3 Patterns across Explored Parameter Ranges

Multidimensional analyses revealed interconnected patterns between epidemiologic and testing parameters while supporting the trends observed in the individual parameter assessments. The best results (lowest cumulative case counts at detection) were obtained in scenarios with high epidemic growth rates (ρ), high testing fractions (π), and lower baseline test counts (μ) across all parameter combinations and models (Figures S2–S5: darkening gradients). This trend was amplified for negative-test-based thresholds (Figure 1), particularly in scenarios with lower proportions of negative tests in the baseline (top rows of heatmaps). The advantage of negative test thresholds, compared to total tests, was thereby largest in scenarios with lower proportions of negative tests—and hence lower negative test baselines. These scenarios exhibited the earliest detections under negative-test thresholds as the lower baselines set a lower detection threshold, which the epidemic’s disproportionately negative test contributions cross more readily.

### 3.4 Epidemic Size Estimates

Epidemic size estimates and widths of corresponding confidence intervals were inversely related to the testing fraction (π) across overdispersion levels (φ=1.0-5.0), baseline test counts (μ=100-1,000), and signal strengths (Figure 2). Across all scenarios, testing fractions below 5% yielded very wide 95% CI that provided very little practical information, while epidemic size estimates and precision were markedly improved for π above 30%.

**Figure 2:**
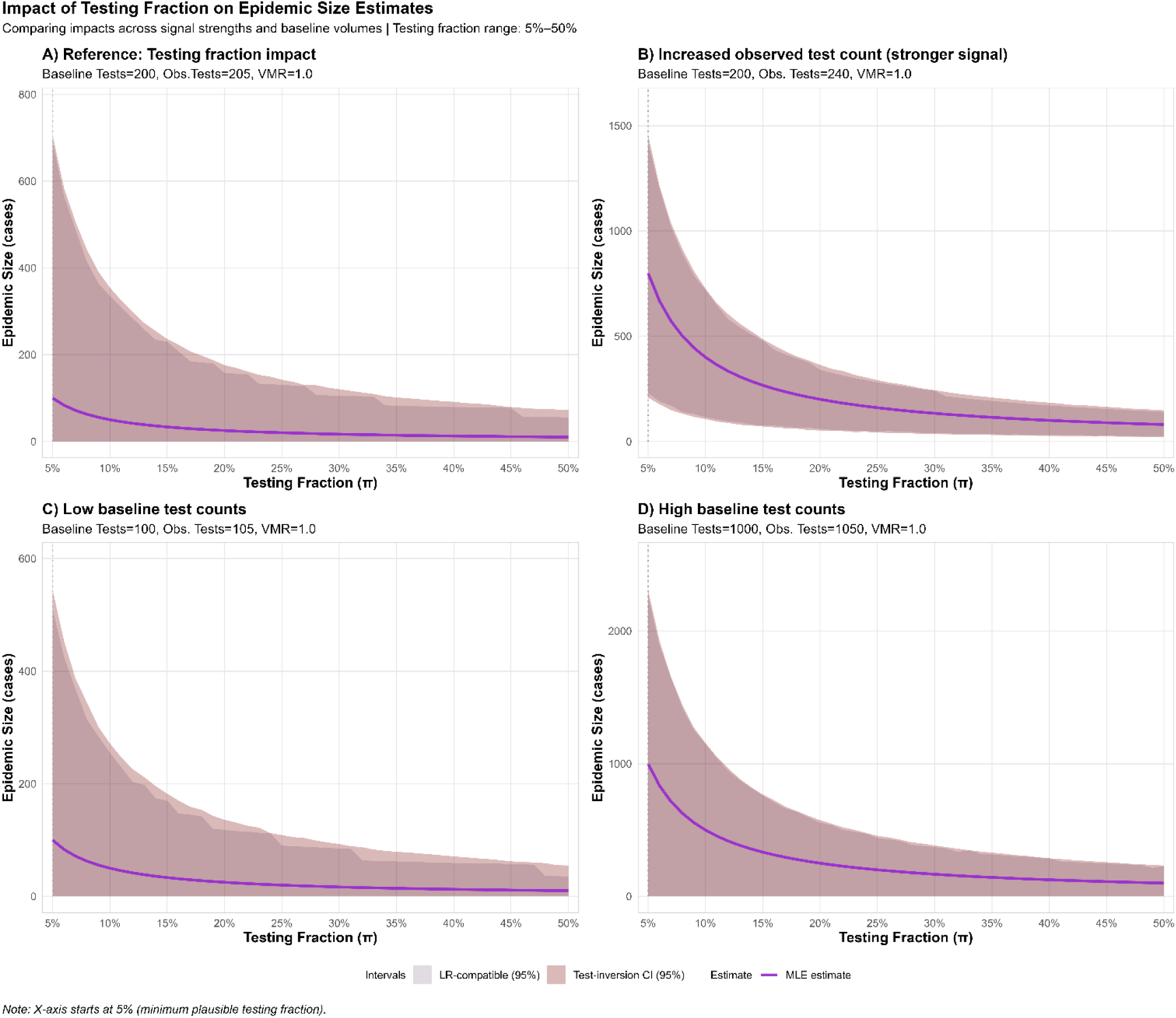
Impact of Testing Fraction on Epidemic Size Estimates and Uncertainty. *Legend*: Estimated size of the underlying epidemic due to the novel ‘mimic’ condition (y-axis, case counts) as a function of testing fractions π (x-axis, 5%–50%), showing an inverse relationship: higher π implies smaller epidemic for fixed observed excess tests. **Solid lines**: Maximum likelihood estimate (MLE) of the underlying epidemic. **Shaded regions**: dark = 95% confidence interval, light = 95% likelihood ratio-compatible set. Panels compare: **(A)** reference case (baseline=200, observed=205); **(B)** stronger signal close to detection threshold (baseline=200, observed=240); **(C)** low baseline volume (baseline=100, observed=105); **(D)** high baseline volume (baseline=1000, observed=1050). All scenarios use variance-to-mean ratio (VMR)=1.0 (Poisson baseline), κ=3 standard deviation threshold. *Note*: X-axes are truncated at 5% (minimum plausible π) to improve readability (low testing fractions π < 0.05 heavily inflate the estimated epidemic size at detection resulting in very large y-axes). The detection approach is not practical for very low testing fractions (π < 0.05) given that only a very small fraction of the underlying emerging epidemic would be captured in the routine testing data.

While absolute uncertainty (95% CI width) decreased with increasing testing fractions, relative uncertainty (95% CI width/ MLE estimate) was primarily driven by signal strength, i.e., the distance between observed tests and threshold (Figure 2 & S6). Relative uncertainty markedly reduced as observed test counts approached the detection threshold (i.e., increasing signal strength, Figure 2B). Uncertainty in the estimated epidemic size was further reduced when observed tests crossed the detection threshold, yielding narrow 95% CI even at low testing fractions (Figure S6). Moreover, baseline test volumes shaped the presentation of uncertainty: low baseline settings yielded small absolute epidemic sizes but large relative uncertainty, whereas high baseline settings generated larger absolute estimates with more stable relative precision (for π=0.1, holding other parameters constant, cf. Figure 2). This scale penalty means that larger surveillance systems establish higher detection thresholds, requiring larger underlying epidemics to trigger detection even at comparable relative precision.

Overdispersion in baseline test counts introduced a multiplicative effect in 95% CI widths, which increased from 1.12 (φ=1.5) to 1.85-times (φ=5.0) compared to the Poisson reference (φ=1.0, cf. Figure 3). Higher overdispersion expanded the detection window based on the same baseline test counts (cf. increasing coverage of the shaded CI along the x-axis in Figure 3). Across all simulations, scenarios where observed test counts were above baseline but clearly below the detection threshold yielded 95% CI that were bound at 0 (Figure 2; ribbons in Figure 3).

**Figure 3:**
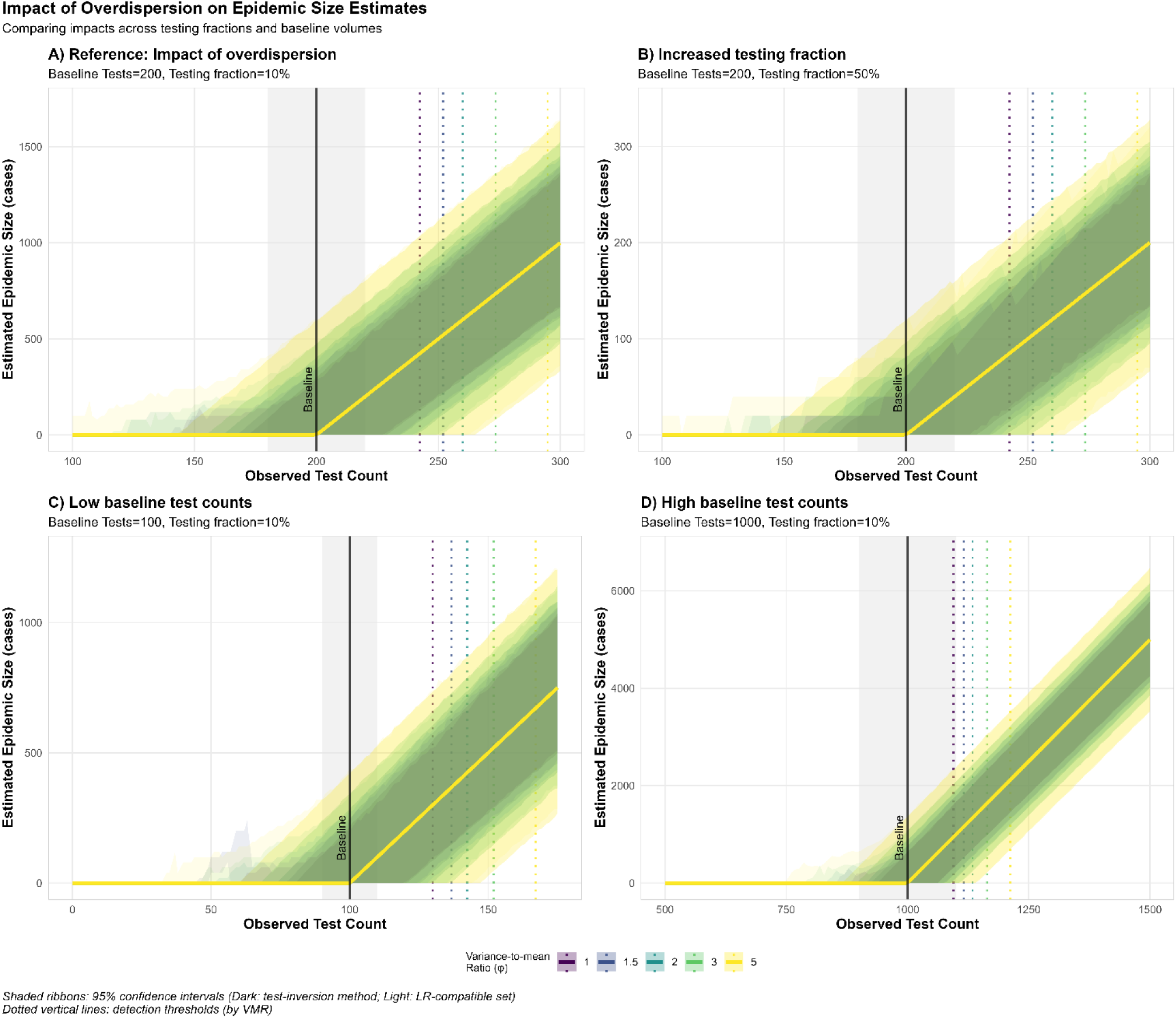
Impact of Baseline Overdispersion on Epidemic Size Estimation Uncertainty. *Legend*: Estimated size of the underlying epidemic due to the novel ‘mimic’ disease (y-axis, case counts) versus observed test counts (x-axis) across variance-to-mean ratios (VMR, φ = 1.0–5.0, colored lines & ribbons). **Solid lines**: Maximum likelihood estimates (MLE). Note that the MLE point estimates derived from the closed-form analytic solution are equivalent for all VMR, so all MLE curves are overlaid (only the curve for φ = 5.0 is visible). **Ribbons**: dark = 95% confidence intervals, light = 95% LR-compatible sets. **Dotted vertical lines**: detection thresholds (κ = 3) colored by VMR. **Solid vertical line**: baseline mean count. **Gray shading**: ±10% baseline range. **Panel A (Reference):** Moderate baseline count (200 tests) with low testing fraction (π = 10%); **(B: Increased Testing Fraction):** Moderate baseline count with high testing fraction (baseline = 200, π = 50%); **(C: Low baseline volume)**: Low baseline counts with low testing fraction (baseline = 100, π = 10%); **(D: High baseline volume)**: High baseline counts with low testing fraction (baseline = 1,000, π = 10%).

In conjunction, these findings underscore that testing fractions (π) and baseline variability (φ) jointly constrain epidemic detection. Bounds on the underlying epidemic size were most informative for signals close to or above the detection threshold; the most actionable, precise intervals occurred under the same stringent conditions that enable early detections: high π and stable, low baseline counts and variability (lower μ and φ).

## 4. Discussion

We evaluated whether routine testing data for a clinically similar (’mimicking’) condition can serve as an early-warning signal for an emerging outbreak, and we show that negative-test counts in particular can provide timely detection under a defined set of surveillance conditions. The approach performs best when the existing test is used infrequently and consistently (low μ, low φ), when novel-disease cases are likely to be tested for the known condition (high π), and when the alert threshold can be tuned to tolerate some false alarms (low κ).

Critically, use of (negative) test results rather than total test volume provides meaningful gains as negative-test thresholds consistently detected outbreaks earlier and at smaller epidemic sizes than total-test approaches while maintaining comparable specificity. However, our findings are cautionary: while the system can be sensitive in favorable settings (high π, low-volume baselines with few negatives), detections are not always reliable and require thousands of epidemic cases in many realistic settings, underscoring that the absence of an alert may not be reassuring. Our framework further showed that estimates of the underlying epidemic size scaled inversely with the testing fraction (1/π), and precision improved as observed tests approached or exceeded the detection threshold. Conversely, larger (negative) baseline counts, and greater variability imposed a scale penalty, slightly delaying alerts and expanding estimate uncertainty. Finally, wide prediction intervals for the number of cases at detection indicate that while the average performance of a system like this may not support its use as a sole means of detecting outbreaks, sometimes it will correctly trigger early outbreak detections, suggesting that as part of a surveillance “mosaic” such systems may be useful as one source of information.

These results align with the literature on statistical outbreak detection and surveillance algorithms, which has long recognized the importance of accurately capturing baseline patterns and overdispersion.^1,3,38,39,42–44^ Our findings extend previous work on alternative surveillance data streams by quantifying conditions under which routine test data provide early outbreak signals, akin to leveraging influenza testing for COVID-19 detection.^17,18,26–28,45,46^ The finding that sensitivity is highest when the emerging pathogen frequently triggers negative tests aligns with the conditions observed during 2022 mpox outbreaks, where presentations often led to negative tests for common STIs.^20,29,30,47^ The cautionary finding that lack of a signal provides limited reassurance—lack of detection remains compatible with substantial undetected epidemics—resonates with established sensitivity-specificity trade-offs in syndromic surveillance^1,15,16^, and underscores the importance of integrating multiple, complementary data streams into surveillance “mosaics”.^21,22,45,48–50^

Our findings suggest how surveillance of tests for conditions easily confused with a novel infection can enhance early epidemic detection, but limitations follow from model simplifications. Baseline test volumes are assumed to be stationary apart from healthcare seeking for the mimicking condition, but may be influenced by disease seasonality, supply chain disruptions, or changes in hospital or provider testing policies, which could mimic or mask signals. Pathogen evolution can likewise degrade assay sensitivity for the mimicking condition, increasing apparent (false) negatives over time and potentially producing signals that resemble—or mask—a novel disease outbreak. While the modeled overdispersion accounts for additional variability, our methodological framework does not specifically account for such complex sources of heterogeneity, which may not be captured in most real-world datasets. Moreover, granular test result data are often unavailable due to privacy concerns, and our frameworks assume conditional independence of testing streams over time, which is violated by temporal correlations arising from day-of-the-week effects, batch processing, or seasonality that can alter detection time distributions and accuracy.

A key strength is the unified quantitative framework combining analytic solutions, stochastic simulations, and likelihood-based estimation across different distributional assumptions and parameter settings. This rigorous approach enables systematic investigation of multidimensional parameter spaces and system dynamics, going beyond assessments based on anecdotal or descriptive evidence. The agreement between analytic and simulation approaches thereby supports the robustness of our results. Importantly, by quantifying the added value of disaggregated negative test data, we provide an evidence-based argument for making such data accessible to enhance public health intelligence.

These findings have direct implications for the design and implementation of public health surveillance. Accurate characterization of baseline variability—not just mean volumes^14^—and stabilizing the monitored data streams (e.g., segmentation or detrending to reduce μ and φ) are critical for reliable detection. Our analyses quantify consequences for monitoring under simplified variance assumptions, which remain relevant for rapid deployment during crises, when expanding to new populations, and in resource-constrained settings. We show that in such high-variability settings, raising testing coverage can yield noticeable returns, supporting prioritization of basic surveillance infrastructure over advanced methods when resources are constrained.

From an operational perspective, public health agencies should prioritize investments in disaggregated test data, as negative test thresholds provided meaningful gains in sensitivity and timeliness. Further, this approach is best targeted towards novel conditions likely to prompt testing (high π) against a background of an infrequently used existing test (low baseline μ). The level of geographic (dis-)aggregation directly shapes both baseline test volume (μ) and variability (φ) and thereby determines favorable detection conditions. Exploratory analysis of NREVSS testing data (Supplement E) supports this relationship: overdispersion is extreme at national scale (φ > 3,000) and high on the level of HHS regions (φ ≈ 91–195), but falls within our modeled range (φ = 1–5) when projected to facility or small-scale catchment areas—the operational scale (μ≈50-500) for which this approach is most plausible. Monitoring at the level of individual facilities, sentinel networks, or geographically defined catchment areas—rather than broad regional or national aggregates—thus provides the most favorable signal-to-noise ratio.

In practice, this favors integration of the approach with existing sentinel surveillance infrastructure where facility-level test results are recorded and baseline characteristics can be estimated directly. For a novel respiratory pathogen, the most practical analog is monitoring influenza-negative ILI specimens within sentinel virologic surveillance or pan-negative results on multiplex respiratory panels against a low-circulation baseline; for a novel STI-mimicking threat, negative syphilis or HSV tests within a clinic network could be leveraged (Table 1).

Validation using real-world data from outbreaks where such mimicking phenomena occurred (e.g., capturing mpox in STI testing, detecting respiratory pathogens in influenza or COVID-19 tests) represents the critical next step. Prospective pilots can guide operational development of adaptive methods to identify optimal detection thresholds that balance timeliness against false alerts for specific data streams and public health contexts. Methodological extensions could incorporate sub-exponential growth, behavioral feedback affecting testing fractions, and temporal dynamics (e.g., seasonality or reporting delays), for example, through state-space or hierarchical models and nowcasting approaches.^51–56^ Long-term goals include investigating adaptation to and integration with alternative data streams,^13,57^ such as syndromic surveillance for chief complaints, and multi-pathogen scenarios^58^ to broaden the flexibility and utility of the framework.

Given the cautionary result that the system is not consistently sensitive, the approach should be treated as a complement to existing surveillance. However, if implementation costs are modest,^59^ the potential for early detection may justify deployment within integrated systems^21,22,37,60,61^ (e.g., sentinel networks or syndromic surveillance).

### 4.1 Conclusion

This study provides a rigorous quantitative foundation for understanding how routine testing streams, especially negative tests, can provide early signals of emerging epidemics under stringent conditions, but in many conditions will not do so until epidemic sizes are sizeable. By elucidating the epidemiologic and operational conditions governing detection, we show the advantage of thresholds based on negative test counts, providing a strong argument for making disaggregated data more accessible for public health surveillance. The approach can be useful in a narrow context with favorable signal-to-noise ratio—it works best for novel conditions that are likely to prompt testing for an existing condition with a stable low-volume baseline. This evidence is thus encouraging for specific use cases but cautionary for broad application; these methods can add value as a low-cost, resource-efficient component within a diverse, multi-source surveillance portfolio, but should not be relied on as stand-alone early warning systems.

## Ethics statement

This study did not require approval from an institutional review board (IRB) or local ethics committee since the research described in this article is solely based on an analytic framework and computational simulations and did not involve any human subjects, human material or personal data.

## Acknowledgements

Anonymized as per journal request

## Supplementary material

Supplementary materials (detailed methods, supplementary tables and figures) are provided in a separate document.

## Contribution statement

**Main author:** Writing – original draft, reviewing & editing, Conceptualization, Methodology, Formal analysis & Validation, Visualization.

**Senior author:** Writing – review & editing, Conceptualization, Methodology, Funding acquisition

## Data availability

No data was used for the research described in the article. The code underlying this manuscript can be made available upon request.

