## Supplementary materials for "Leveraging Limited Testing Data for Early Detection of Emerging Infectious Disease Outbreaks"

**List of included materials:**

Key parameter definitions

Supplementary methods

A: Comprehensive Table for Novel Infection – Mimicking Condition Pairs

B: Closed-form Solutions under Poisson Assumptions

C: Convolution Approach for Negative Binomial Baselines

D: Detailed Methods for Estimating the Underlying Epidemic Size from Routine Testing Data

E: Empirical assessment of overdispersion in surveillance data

F: Stochastic Simulation Methods

Supplementary results

Tables S1 & S2

Figures S1 – S6

Alt text for figures

Used R packages

**Key parameter definitions**

μ: Mean (expected) count of baseline tests per time unit (positive or negative)

> *Note*: For consistency with the mathematical literature, the mean (expected) count of baseline tests per time unit in Poisson models (Supplement B) is defined as λ

$\sigma^2$: Variance of (expected) routine test counts (positive or negative) (Poisson: $\sigma^2 = \lambda$)

φ: Variance-to-mean ratio of baseline tests (positive or negative)

π: Fraction of epidemic cases that undergoes routine testing for the known condition; probability of getting tested for the routinely monitored known condition among epidemic cases per t

ρ: Epidemic growth rate (exponential) of the unknown disease per t

κ: Detection threshold sensitivity parameter balancing early detections with false positives

τ: Time of epidemic emergence

*f*: Fraction of total tests that are routinely negative

$\widehat{N}_{Epi_{MLE}}$: Maximum likelihood estimate (MLE) of the underlying epidemic

**Supplementary methods**

**Supplement A: Comprehensive Table for Novel Infection – Mimicking Condition Pairs with Supporting References**

**Overview:** This table provides documented examples of emerging or novel infections that clinically mimic established conditions for which routine diagnostic testing exists. These pairs represent potential use cases for the test-volume-based detection framework described in the manuscript.

---

#### 1. Mpox → Sexually transmitted infections (e.g., syphilis, HSV)

**Mimicry mechanism:** Mpox genital/anogenital lesions closely resemble primary syphilis chancres and herpes simplex vesicles. During the 2022 global outbreak, many mpox patients were initially evaluated and tested for STIs, with 15-28% having concurrent STI diagnoses and many testing negative for the STI they were initially suspected of having.

**Surveillance data stream:** STI testing data (syphilis serology, HSV PCR, gonorrhea/chlamydia NAAT); clinic-level or jurisdiction-level STI testing volumes.

| # | Reference | Key relevance |
|---|---|---|
| 1 | Thornhill JP, Barkati S, Walmsley S, et al. Monkeypox Virus Infection in Humans across 16 Countries – April-June 2022. *N Engl J Med*. 2022;387(8):679-691. *doi:10.1056/NEJMoa2207323* | Largest early multinational case series (528 cases); documented clinical overlap with STIs and frequent concurrent STI testing |
| 2 | Ciccarese G, Di Biagio A, Drago F, et al. Monkeypox virus infection mimicking primary syphilis. *Infez Med*. 2023;31(1):113-115. *doi:10.53854/liim-3101-16* | Direct documentation of mpox lesions indistinguishable from primary syphilis chancre |
| 3 | Long B, Liang SY, Carius BM, et al. Mimics of Monkeypox: Considerations for the emergency medicine clinician. *Am J Emerg Med*. 2023;65:172-178. *doi:10.1016/j.ajem.2023.01.007* | Comprehensive review that directly supports the central thesis of the manuscript by describing mpox mimics including syphilis, HSV, and other STIs. |
| 4 | Gunaratne S, Vigil K, Huang S et al. STI testing and diagnosis rates among patients with Mpox in 2 populous US cities, 2022. *BMC Medicine*. 2025. *doi:10.1186/s12916-025-04396-1* | 66% of mpox patients underwent STI testing; 17.5% diagnosed with concurrent STIs, syphilis most common |
| 5 | Mourad A, Alavian N, Woodhouse E et al. Concurrent Sexually Transmitted Infection Testing Among Patients Tested for Mpox at a Tertiary Healthcare System. *Open Forum Infect Dis*. 2023;10(8):ofad381. *doi:10.1093/ofid/ofad381* | 15.2% concurrent STI diagnosis rate among mpox-tested patients |
| 6 | Allan-Blitz LT, Gandhi M, Adamson P, et al. A Position Statement on Mpox as a Sexually Transmitted Disease, *Clinical Infectious Diseases*, Volume 76, Issue 8, 15 April 2023, Pages 1508– | Directly highlights and provides comprehensive evidence for the clinical overlap between mpox and STIs |

| # | Reference | Key relevance |
|---|---|---|
| | 1512, *https://doi-org.ezp-prod1.hul.harvard.edu/10.1093/cid/ciac960* | |
| 7 | Curran KG, Eberly K, Russell O et al. HIV and Sexually Transmitted Infections Among Persons with Monkeypox — Eight U.S. Jurisdictions, May 17–July 22, 2022. *MMWR Morb Mortal Wkly Rep*. 2022;71. *doi:10.15585/mmwr.mm7136a1* | Highlights the overlap between mpox and common STIs; argues that evaluation for mpox should be considered in screening for HIV and other STIs |

#### 2. Pandemic influenza → Seasonal influenza

**Mimicry mechanism:** Novel pandemic influenza strains may present identically or very similar to seasonal influenza but test negative for circulating seasonal subtypes on standard subtyping assays. The 2009 H1N1 pandemic was first identified when specimens from ILI cases in southern California tested positive for influenza A but could not be subtyped (i.e., were negative for seasonal H1N1 and H3N2). In Mexico, unusually severe ILI clusters preceded official recognition.

**Surveillance data stream:** Influenza virologic surveillance (subtyping data from sentinel sites, public health laboratories); "unsubtypable" influenza A specimens; ILI syndromic surveillance.

| # | Reference | Key relevance |
|---|---|---|
| 1 | Dawood FS, Jain S, Finelli L, et al. Emergence of a Novel Swine-Origin Influenza A (H1N1) Virus in Humans. *N Engl J Med*. 2009;360(25):2605-2615. *doi:10.1056/NEJMoa0903810* | Definitive account of initial detection: two children in southern California with influenza A positive but unsubtypable by standard assays |
| 2 | CDC. Outbreak of Swine-Origin Influenza A (H1N1) Virus Infection – Mexico, March-April 2009. *MMWR*. 2009;58(17):467-470 | Official CDC report documenting the Mexican outbreak cluster detected through ILI surveillance |
| 3 | Chowell G, Echevarria-Zuno S, Viboud C, et al. Characterizing the Epidemiology of the 2009 Influenza A/H1N1 Pandemic in Mexico. *PLoS Med*. 2011;8(5):e1000436. *doi:10.1371/journal.pmed.1000436* | Retrospective analysis showing early pandemic signals in Mexican surveillance data before official recognition |
| 4 | Hsieh YH, Ma S, Velasco-Hernandez J et al. Early Outbreak of 2009 Influenza A (H1N1) in Mexico Prior to Identification of pH1N1 Virus. *PLoS One*. 2011;7(8):e23853. *doi:10.1371/journal.pone.0023853* | Identified February-March 2009 outbreak in Mexico via excess influenza activity in surveillance data |
| 5 | Zepeda-Lopez HM, Perea-Araujo L, Miliar-Garcia A, et al. Inside the Outbreak of the 2009 Influenza A (H1N1)v Virus in Mexico. *PLoS One*. 2010;5(10):e13256. *doi:10.1371/journal.pone.0013256* | Detailed documentation of 202 confirmed H1N1 cases and detection pathway in Mexico City |
| 6 | Honigsbaum M. Revisiting the 1957 and 1968 influenza pandemics. *Lancet*. 2020;395(10225):621-628. *doi:10.1016/S0140-6736(19)33065-1* | Historical context: novel H2N2 (1957) and H3N2 (1968) pandemic strains were also initially detected through routine influenza surveillance |
| 7 | Taubenberger JK, Morens DM. Influenza: the once and future pandemic. Public Health Rep. 2010 Apr;125 Suppl 3(Suppl 3):16-26. PMID: 20568566; PMCID: PMC2862331. | Provides a historical overview over past influenza pandemics, highlighting that 20th/21st century pandemics were detected via seasonal influenza surveillance. |

#### 3. COVID-19 → Influenza / ILI surveillance

**Mimicry mechanism:** SARS-CoV-2 causes influenza-like illness (fever, cough, myalgia) and was circulating undetected while patients sought care and were tested for influenza. A surge in "influenza-negative ILI" was detectable in routine surveillance data in multiple countries weeks before COVID-19 was officially recognized. The Seattle Flu Study famously identified the first US community transmission case by testing stored influenza surveillance specimens for SARS-CoV-2.

**Surveillance data stream:** Influenza sentinel surveillance (ILI rates, influenza test positivity/negativity); influenza-negative ILI as a specific signal; multiplex respiratory pathogen panel data.

| # | Reference | Key relevance |
|---|---|---|
| 1 | Chu HY, Englund JA, Starita LM, et al. Early Detection of Covid-19 through a Citywide Pandemic Surveillance Platform. *N Engl J Med*. 2020;383:185-187. *doi:10.1056/NEJMc2008646* | Seattle Flu Study: first documented US community transmission (Feb 24, 2020) detected through an influenza surveillance platform |
| 2 | Silverman JD, Hupert N, Washburne AD. Using influenza surveillance networks to estimate state-specific prevalence of SARS-CoV-2 in the United States. *Sci Transl Med*. 2020;12(554):eabc1126. *doi:10.1126/scitranslmed.abc1126* | Used non-influenza ILI surge to estimate 8.7 million SARS-CoV-2 infections in US during March 2020 – direct demonstration of the "test-negative" signal concept; underscores high numbers of tests needed to detect outbreaks using the "test-negative" signal approach |
| 3 | Cobb NL, Collier S, Attia EF et al. Global influenza surveillance systems to detect the spread of influenza-negative influenza-like illness during the COVID-19 pandemic: Time series outlier analyses from 2015-2020. *PLoS Med*. 2022;19(7):e1004035. *doi:10.1371/journal.pmed.1004035* | Influenza-negative ILI outliers predated COVID-19 peaks by an average of 13.3 weeks in 16 of 28 countries studied |
| 4 | Lu FS, Nguyen AT, Link NB, et al. Estimating the cumulative incidence of COVID-19 in the United States using influenza surveillance, virologic testing, and mortality data. *PLoS Comput Biol*. 2021;17(6):e1008994. *doi:10.1371/journal.pcbi.1008994* | Four complementary approaches using influenza surveillance data to estimate true COVID-19 incidence |
| 5 | Wen A, Wang L, He H, et al. An aberration detection-based approach for sentinel syndromic surveillance of COVID-19 and other novel influenza-like illnesses. *J Biomed Inform*. 2021;113:103660. *doi:10.1016/j.jbi.2020.103660* | Demonstrated that aberration detection algorithms applied to ILI data could have provided early COVID-19 warning |
| 6 | Marcenac P, McCarron M, Davis W et al. Leveraging International Influenza Surveillance Systems and Programs during the COVID-19 Pandemic. *Emerg Infect Dis*. | 82% of CDC Influenza Division partner countries adapted existing influenza surveillance for COVID-19 detection by May 2020 |

| # | Reference | Key relevance |
|---|---|---|
| | 2022;28(13 Suppl). *doi:10.3201/eid2813.212248* | |
| 7 | Kogan NE, Clemente L, Liautaud P et al. An early warning approach to monitor COVID-19 activity with multiple digital traces in near real time.*Sci.Adv.***7**,eabd6989(2021).DOI:*10.1126/sciadv.abd6989* | Assesses ILI surveillance and virological influenza surveillance data as early indicators of COVID-19 activity |

#### 4. Emerging neurotropic pathogens → Polio / AFP surveillance

**Mimicry mechanism:** Acute flaccid paralysis (AFP) surveillance, established globally for polio eradication, captures all cases of acute limb weakness regardless of etiology. Non-polio enteroviruses (especially EV-D68 and EV-A71), West Nile virus, and other neurotropic pathogens cause polio-like acute flaccid myelitis (AFM) that enters the AFP surveillance pipeline and tests negative for poliovirus. The 2014 and 2018 EV-D68 outbreaks in the US were detected through this mechanism.

**Surveillance data stream:** AFP case reports and poliovirus testing results from global polio surveillance infrastructure; "non-polio AFP" rates.

| # | Reference | Key relevance |
|---|---|---|
| 1 | Messacar K, Asturias EJ, Hixon AM, et al. Enterovirus D68 and acute flaccid myelitis – evaluating the evidence for causality. *Lancet Infect Dis*. 2018;18(8):e239-e247. *doi:10.1016/S1473-3099(18)30094-X* | Systematic evaluation of EV-D68 as cause of polio-like AFM using Bradford Hill criteria |
| 2 | Ayscue P, Van Haren K, Glaser C, et al. Acute Flaccid Paralysis with Anterior Myelitis – California, June 2012-June 2014. *MMWR*. 2014;63(40):903-906 | Early identification of AFM cluster through AFP surveillance; EV-D68 detected in respiratory specimens |
| 3 | Carmona RCC, Reis FC, Cilli A et al. Beyond Poliomyelitis: A 21-Year Study of Non-Polio Enterovirus Genotyping and Its Relevance in Acute Flaccid Paralysis in Sao Paulo, Brazil. *Viruses*. 2024;16(12):1875. *doi:10.3390/v16121875* | 21-year study showing non-polio enteroviruses in 6.9% of AFP cases – demonstrates AFP surveillance capturing non-polio neurotropic pathogens |
| 4 | Sejvar JJ, Leis AA, Stokic DS et al. Acute Flaccid Paralysis and West Nile Virus Infection. *Emerg Infect Dis*. 2003;9(7):763-769. *doi:10.3201/eid0907.030129* | West Nile virus causing poliomyelitis-like syndrome detected through AFP-like surveillance |
| 5 | WHO Global Polio Eradication Initiative. Global Guidelines for Acute Flaccid Paralysis (AFP) Surveillance. 2023. Available: *https://polioeradication.org* | Official WHO framework describing AFP surveillance infrastructure and its capacity for detecting non-polio neurotropic pathogens |
| 6 | Messacar K, Matzinger S, Berg K et al. Multimodal Surveillance Model for Enterovirus D68 Respiratory Disease and Acute Flaccid Myelitis among Children in Colorado, USA, 2022. *Emerg Infect Dis*. 2024;30(3):544-555. *doi:10.3201/eid3003.231223* | Modern multimodal surveillance integrating EV-D68 respiratory and AFM detection (including wastewater surveillance developed for polio) |

#### 5. Smallpox (or novel poxviruses) → Chickenpox (varicella)

**Mimicry mechanism:** Smallpox (variola) produces a vesicular/pustular rash that can be confused with chickenpox (varicella), particularly by clinicians with no experience of smallpox (eradicated 1980). In a bioterrorism or natural re-emergence scenario, early cases would likely be initially diagnosed as chickenpox. Mpox is also frequently confused with varicella in African endemic settings, with co-infection rates of ~12% documented in the DRC.

**Surveillance data stream:** Varicella surveillance data; vesicular rash illness reporting; poxvirus-negative varicella testing.

| # | Reference | Key relevance |
|---|---|---|
| 1 | Henderson DA, Inglesby TV, Bartlett JG, et al. Smallpox as a Biological Weapon: Medical and Public Health Management. *JAMA*. 1999;281(22):2127-2137. *doi:10.1001/jama.281.22.2127* | Consensus statement explicitly discussing smallpox-chickenpox differential diagnosis challenges and the risk of delayed recognition |
| 2 | Breman JG, Henderson DA. Diagnosis and Management of Smallpox. *N Engl J Med*. 2002;346(17):1300-1308. *doi:10.1056/NEJMra020025* | Clinical guidance on distinguishing smallpox from chickenpox (centrifugal vs. centripetal distribution, lesion synchrony) |
| 3 | CDC. Evaluating Patients for Smallpox: Acute, Generalized Vesicular or Pustular Rash Illness Protocol. 2024. Available: *https://www.cdc.gov/smallpox/hcp/diagnosis-testing/* | Current CDC diagnostic algorithm for triaging vesicular rash illness (smallpox vs. chickenpox vs. other) |
| 4 | Seguin D & Stoner Halpern J. Triage of a Febrile Patient with a Rash: A Comparison of Chickenpox, Monkeypox, and Smallpox. *Emerg Infect Dis*. 2022;28(9):1818-1824. PMC9533829 | Direct three-way clinical comparison of poxvirus presentations for differential diagnosis |
| 5 | Jezek Z, Szczeniowski M, Paluku K et al. Human Monkeypox: Confusion with Chickenpox. *Lancet*. 1989. PMID: 2907258 | Early documentation of mpox/varicella diagnostic confusion in endemic settings |
| 6 | Tayachew A, Kebede N, Alayu M et al. Genomic Evidence of Varicella-Zoster Virus Among Mpox-Suspected Cases in Ethiopia During the 2022 Mpox Multi-Country Outbreak. *Sci Rep*. 2025;15:2931. *doi:10.1038/s41598-025-29116-w* | 80.5% of mpox-suspected cases in Ethiopia were misdiagnosed varicella cases – demonstrates bidirectional misclassification. |
| 7 | Hughes, CM, Liu L, Davidson WB et al. A Tale of Two Viruses: Coinfections of Monkeypox and Varicella Zoster Virus in the Democratic Republic of Congo. *Am J Trop Med Hyg*. 2020;104(2):604-611. Published 2020 Dec 7. *doi:10.4269/ajtmh.20-0589* | Highlights challenges in clinical diagnosis of mpox based on rash examination and provides comprehensive overview of similarities and differences between mpox and varicella. |
| 8 | Long B, Liang SY, Carius BM, et al. Mimics of Monkeypox: Considerations for the emergency medicine clinician. *Am J Emerg Med*. | Describes mimics of monkeypox, including smallpox, varicella, syphilis, acute retroviral syndrome, and HSV. |

| # | Reference | Key relevance |
|---|---|---|
| | 2023;65:172-178. *doi:10.1016/j.ajem.2023.01.007* | |

#### 6. Bioterrorism agents → Common endemic conditions

**Mimicry mechanism:** Category A bioterrorism agents frequently present clinically similar to common conditions (esp. in initial stages): inhalational anthrax mimics influenza/community-acquired pneumonia; plague mimics severe pneumonia; botulism mimics descending paralysis/neurological conditions; VHFs mimic severe febrile illness/malaria. Increasing recognition of this clinical overlap following 9/11 drove the development of syndromic surveillance in the early 2000s.

**Surveillance data stream:** Syndromic surveillance (ED chief complaints, ILI reporting, pneumonia surveillance); laboratory testing for common conditions where unexpected negatives might signal a novel/deliberate agent.

| # | Reference | Key relevance |
|---|---|---|
| 1 | Inglesby TV, O'Toole T, Henderson DA, et al. Anthrax as a Biological Weapon, 2002: Updated Recommendations for Management. *JAMA*. 2002;287(17):2236-2252. PMID: 11980524 | Consensus statement documenting anthrax mimicking influenza and pneumonia; foundational for syndromic surveillance rationale |
| 2 | Inglesby TV, Dennis DT, Henderson DA, et al. Plague as a Biological Weapon. *JAMA*. 2000;283(17):2281-2290. PMID: 10807389 | Pneumonic plague initially indistinguishable from severe CAP; detection requires high clinical suspicion |
| 3 | Borio L, Inglesby T, Peters CJ, et al. Hemorrhagic Fever Viruses as Biological Weapons: Medical and Public Health Management. *JAMA*. 2002;287(18):2391-2405. PMID: 11988060 | VHFs presenting as non-specific febrile illness; differential includes malaria, typhoid, and other endemic fevers |
| 4 | Henning KJ. Overview of Syndromic Surveillance. What is Syndromic Surveillance? *MMWR Suppl*. 2004;53:5-11. Available: *https://www.cdc.gov/mmwr/preview/mmwrhtml/su5301a3.htm* | Foundational overview of syndromic surveillance, motivated explicitly as a bioterrorism early warning and detection tool |
| 5 | Buehler JW, Berkelman RL, Hartley DM, Peters CJ et al. Syndromic Surveillance and Bioterrorism-related Epidemics. *Emerg Infect Dis*. 2003;9(10):1197-1204. PMID: 14609452 | Early evaluation of syndromic surveillance performance for bioterrorism detection |
| 6 | Dennis DT, Inglesby TV, Henderson DA, et al. Tularemia as a Biological Weapon: Medical and Public Health Management. *JAMA.* 2001;285(21):2763–2773. *doi:10.1001/jama.285.21.2763* | Highlights difficulty of distinguishing tularemia from community-acquired pneumonia and overlap with other potential bioterrorism agents like plague, anthrax, or Q fever |

#### 7. Nipah virus → Japanese encephalitis / endemic encephalitis

**Mimicry mechanism:** Nipah virus encephalitis presents with fever, headache, altered consciousness, and seizures – clinically indistinguishable from Japanese encephalitis (JE) in endemic regions. The 1998-1999 Malaysia outbreak (265 cases, 105 deaths) was initially misdiagnosed as JE based on clinical presentation and cross-reactive serology. In Bangladesh, Nipah cases are detected through hospital-based encephalitis sentinel surveillance, where they must be differentiated from JE, cerebral malaria, and other endemic diseases.

**Surveillance data stream:** Encephalitis sentinel surveillance; JE testing data (Nipah cases test negative for JE); AFP/acute encephalitis syndrome (AES) surveillance in South and Southeast Asia.

| # | Reference | Key relevance |
|---|---|---|
| 1 | Chua KB, Goh KJ, Wong KT, et al. Clinical Features of Nipah Virus Encephalitis among Pig Farmers in Malaysia. *N Engl J Med*. 2000;342(17):1229-1235. *doi:10.1056/NEJM200004273421701* | First documented outbreak: 265 cases initially diagnosed as JE; only recognized as novel paramyxovirus due to different clinical and epidemiologic features and as ¾ of cases reported JE immunization |
| 2 | Lim CC, Sitoh YY, Hui F, et al. Nipah viral encephalitis or Japanese encephalitis? MR findings in a new zoonotic disease. *AJNR Am J Neuroradiol*. 2000;21(3):455-461. PMID: 10730635 | MRI differentiation between Nipah and JE encephalitis; demonstrates initial diagnostic confusion; all cases tested negative for JE |
| 3 | CDC. Outbreak of Hendra-Like Virus – Malaysia and Singapore, 1998-1999. *MMWR*. 1999. Available: *https://www.cdc.gov/mmwr/preview/mmwrhtml/00056866.htm* | Official CDC report documenting initial JE misdiagnosis and subsequent identification of Nipah as "Hendra-like virus" |
| 4 | Satter SM, Aquib WR, Sultana S et al. Tackling a global epidemic threat: Nipah surveillance in Bangladesh, 2006-2021. *PLoS Negl Trop Dis*. 2023;17(9):e0011617. *doi:10.1371/journal.pntd.0011617* | 15-year hospital-based encephalitis sentinel surveillance detecting 322 Nipah cases (71% CFR) from suspected meningo-encephalitis cases; built in analogy to JE surveillance |
| 5 | Naser AM, Hossain MJ, Sazzad HMS et al. Integrated cluster- and case-based surveillance for detecting stage III zoonotic pathogens: an example of Nipah virus surveillance in Bangladesh. Epidemiol Infect. 2015 Jul;143(9):1922-30. doi: 10.1017/S0950268814002635 | Integrated surveillance for Nipah virus based on encephalitis surveillance infrastructure |
| 6 | Annand EJ, Horsburgh BA, Xu K et al. Novel Hendra Virus Variant Detected by Sentinel Surveillance of Horses in Australia. *Emerg Infect Dis*. 2022;28(3):648-652. *doi:10.3201/eid2813.211245* | Related paramyxovirus (Hendra) detected through equine sentinel surveillance; novel variant missed by standard PCR |

#### 8. Novel pandemic respiratory pathogen → Existing respiratory virus testing (COVID-19, influenza, RSV)

**Mimicry mechanism:** A future novel respiratory pathogen would present as ILI/ARI and be tested using existing multiplex panels for SARS-CoV-2, influenza, and RSV. Testing negative for all known targets diseases could serve as a signal. The WHO Mosaic Surveillance Framework explicitly envisions integrating multiple respiratory data streams for pathogen-agnostic detection – this framework could be used for the prospective application of the detection approach presented in the current study.

**Surveillance data stream:** Multiplex respiratory pathogen panel data (negative for all target diseases); sentinel ILI/ARI surveillance with virologic testing; wastewater surveillance for respiratory viruses.

| # | Reference | Key relevance |
|---|---|---|
| 1 | Mott JA, Bergeri I, Lewis HC, et al. Facing the future of respiratory virus surveillance: "The mosaic surveillance framework." *Influenza Other Respir Viruses*. 2023;17(3):e13122. *doi:10.1111/irv.13122* | Brief description of the WHO framework (see 2 below) for integrating multiple respiratory surveillance streams, including pathogen-agnostic detection. Presents a systematic framework for the approach presented in our study. Highlights commonalities of respiratory viruses. |
| 2 | WHO. Crafting the mosaic: a framework for resilient surveillance for respiratory viruses of epidemic and pandemic potential. 2023. Available: *https://www.who.int/publications/i/item/9789240070288* | Full WHO guidance document on mosaic surveillance design for pandemic preparedness |
| 3 | Cheemarla NR, Hanron A, Fauver JR, et al. Nasal host response-based screening for undiagnosed respiratory viruses: a pathogen surveillance and detection study. *Lancet Microbe*. 2023;4(1):e38-e46. *doi:10.1016/S2666-5247(22)00296-8* | Novel approach using host transcriptomic response to detect undiagnosed respiratory infections in test-negative specimens. Highlights additional biomarkers (host and pathogen) to make detection approach more practical. |
| 4 | Jules E, Decker C, Bixler BJ, et al. Respiratory Virus Detection and Sequencing from SARS-CoV-2-Negative Rapid Antigen Tests. *Emerg Infect Dis*. 2025;31(13):39-44. *doi:10.3201/eid3113.241191* | Demonstrates that SARS-CoV-2-negative rapid tests can be repurposed for detecting other respiratory viruses with overlapping symptoms |
| 5 | Gur-Arie L, Stein M, Sefty H, et al. Hospital surveillance of respiratory viruses during the COVID-19 pandemic and beyond: contribution to the WHO mosaic framework, Israel, 2020 to 2023. *Eurosurveillance*. 2023;28(40):2300528. *doi:10.2807/1560-7917.ES.2023.28.40.2300528* | Real-world implementation of integrated respiratory virus surveillance within the mosaic framework |
| 6 | Boehm AB, Hughes B, Duong D, et al. Wastewater concentrations of human influenza, metapneumovirus, parainfluenza, respiratory syncytial virus, rhinovirus, and | Multi-pathogen wastewater surveillance as complementary data stream for respiratory pathogen detection. |

| # | Reference | Key relevance |
|---|---|---|
| | seasonal coronavirus nucleic-acids during the COVID-19 pandemic. *Lancet Microbe*. 2023;4(5):e340-e348. *doi:10.1016/S2666-5247(22)00386-X* | |
| 7 | Marcenac P, McCarron M, Davis W, et al. Leveraging International Influenza Surveillance Systems and Programs during the COVID-19 Pandemic. *Emerg Infect Dis*. 2022;28(13):S26-S33. *doi:10.3201/eid2813.212248* | Describes how countries used existing surveillance systems for severe acute respiratory infection and influenza-like illness, respiratory virus laboratory resources, and ongoing population-based influenza studies to track, study, and respond to SARS-CoV-2 infections. Highlights how incorporation of COVID-19 surveillance into existing influenza sentinel surveillance systems can support continued global surveillance for respiratory viruses with pandemic potential |

9. Zika virus → Dengue fever

**Mimicry mechanism:** Zika virus causes a dengue-like febrile illness with rash, arthralgia, and conjunctivitis. During the 2015-2016 epidemic in the Americas, Zika was initially detected through dengue surveillance systems when patients presenting with dengue-like illness tested negative for dengue. Serological cross-reactivity between flaviviruses further complicates diagnosis. In Brazil, the Zika outbreak in Bahia was identified when specimens from suspected dengue patients tested negative for DENV, CHIKV, and other arboviruses.

**Surveillance data stream:** Dengue surveillance (syndromic + virologic); arboviral multiplex testing panels; dengue-negative febrile illness rates.

| # | Reference | Key relevance |
|---|---|---|
| 1 | Campos GS, Bandeira AC, Sardi SI. Zika Virus Outbreak, Bahia, Brazil. *Emerg Infect Dis*. 2015;21(10):1885-1886. *doi:10.3201/eid2110.150847* | First identification of Zika in Brazil through testing of suspected dengue cases; samples tested negative for DENV and CHIKV before ZIKV was detected. Also mentions identification of ZIKV via yellow fever surveillance |
| 2 | Brasil P, Pereira JP Jr, Moreira ME, et al. Zika Virus Outbreak in Rio de Janeiro, Brazil: Clinical Characterization, Epidemiological and Virological Aspects. *PLoS Negl Trop Dis*. 2016;10(4):e0004636. *doi:10.1371/journal.pntd.0004636* | Highlights clinical characterization showing dengue-like presentation; detection through arboviral surveillance (focused on “unusual dengue”) |
| 3 | Musso D, Ko AI, Baud D. Zika Virus Infection – After the Pandemic. *N Engl J Med*. 2019;381:1444-1457. *doi:10.1056/NEJMra1808246* | Comprehensive review including initial detection of Zika outbreaks through dengue surveillance infrastructure |
| 4 | Sharp TM, Ryff KR, Alvarado L et al. Surveillance for Chikungunya and Dengue During the First Year of Chikungunya Virus Circulation in Puerto Rico. *J Infect Dis*. 2016;214(S5):S475-S481. *doi:10.1093/infdis/jiw245* | Demonstrates arboviral surveillance system detecting novel pathogen (CHIKV) against cocomitant dengue circulation |
| 5 | Rico-Mendoza A, Porras-Ramirex A, Chang A et al. Co-circulation of dengue, chikungunya, and Zika viruses in Colombia from 2008 to 2018. *Rev Panam Salud Publica*. 2019;43:e48. *doi:10.26633/RPSP.2019.48* | Describes co-circulation of multiple arboviral pathogens with similar clinical presentation complicating surveillance; highlights that multiplex approaches are needed |

#### 10. Ebola / VHFs → Malaria and other endemic febrile illness

**Mimicry mechanism:** Ebola virus disease (EVD) initially presents as non-specific febrile illness (fever, headache, myalgia, fatigue) that is clinically indistinguishable from malaria, typhoid fever, and other common tropical febrile illnesses. During the 2013-2016 West African epidemic, early Ebola cases were initially treated as malaria, delaying recognition. Conversely, malaria diagnosis was disrupted because patients avoided healthcare facilities, and Ebola screening protocols sometimes missed concurrent malaria.

**Surveillance data stream:** Malaria rapid diagnostic test (RDT) data; febrile illness surveillance; syndromic surveillance for hemorrhagic fever.

| # | Reference | Key relevance |
|---|---|---|
| 1 | Borio L, Inglesby T, Peters CJ, et al. Hemorrhagic Fever Viruses as Biological Weapons: Medical and Public Health Management. *JAMA*. 2002;287(18):2391-2405. PMID: 11988060 | VHFs presenting as non-specific febrile illness; differential diagnosis includes malaria, dengue and typhoid. Also highlights lack of widely available diagnostic tests for VHF |
| 2 | Boggild AK, Esposito DH, Kozarsky PE Differential diagnosis of illness in travelers arriving from Sierra Leone, Liberia, or Guinea: A Cross-sectional Study From the GeoSentinel Surveillance Network. *Ann Intern Med*. 2015;162(11):757-764. *doi:10.7326/M15-0074* | GeoSentinel data showing malaria as most common diagnosis among travelers from Ebola-affected countries during the 2013-2016 epidemic |
| 3 | Chua AC, Cunningham J, Moussy F et al. The Case for Improved Diagnostic Tools to Control Ebola Virus Disease in West Africa and How to Get There. *PLoS Negl Trop Dis*. 2015;9(4):e0003734. *doi:10.1371/journal.pntd.0003734* | Describes diagnostic challenges distinguishing EVD from malaria and other endemic febrile illness. Underscores barriers to decentralized detection of EVD in resource-constrained settings |
| 4 | Tan KR, Cullen KA, Koumans EH & Arguin PM. Inadequate Diagnosis and Treatment of Malaria Among Travelers Returning from Africa During the Ebola Epidemic – United States, 2014-2015. *MMWR*. 2016;65(5):105-107. *doi:10.15585/mmwr.mm6505a3* | Highlights clinical overlap via documenting diagnostic disruption: Ebola concerns interfered with malaria diagnosis among returning travelers |
| 5 | Zalwango, J.F., Naiga, H.N., Nsubuga, E.J. *et al.* Understanding the delay in identifying Sudan Virus Disease: gaps in integrated disease surveillance and response and community-based surveillance to detect viral hemorrhagic fever outbreaks in Uganda, September 2022. *BMC Infect Dis* **24**, 754 (2024). *https://doi-org.ezp-prod1.hul.harvard.edu/10.1186/s12879-024-09659-5* | Highlights similar clinical presentation of Ebola and more prevalent malaria and typhoid fever, creating challenges in detecting Ebola disease outbreaks |

**Supplement B: Closed-form Solutions under Poisson Assumptions**

When baseline test count variability follows a Poisson process ($\varphi = 1$, i.e., $\sigma^2 = \mu$), the detection framework can be represented via closed-form analytic solutions. This special case serves as a useful benchmark for the lower bounds of detection and provides mathematical insight into the detection mechanism.

**B1. Detection threshold**

Under Poisson distributional assumptions, the baseline threshold simplifies to:

$$\mathfrak{T} = \lambda_{\text{Base}} + \kappa\sqrt{\lambda_{\text{Base}}} \quad \text{(Eq. B1)}$$

where $\lambda_{\text{Base}} = \lambda_{\text{Pos}} + \lambda_{\text{Neg}}$ is the expected total baseline test count, and the standard deviation is $\sqrt{\lambda_{\text{Base}}}$ by the Poisson variance property.

For negative test thresholds:

$$\mathfrak{T}_{Neg} = \lambda_{\text{Neg}} + \kappa\sqrt{\lambda_{\text{Neg}}} \quad \text{(Eq. B2)}$$

**B2. Detection probability**

After epidemic emergence at $\tau$, the total expected test count at time $t$ becomes:

$$\lambda_{\text{Tot}}(t) = \underbrace{\pi \cdot \exp(\rho[t-\tau])}_{Epidemic-driven\ tests} + \lambda_{\text{Base}}$$

Since the sum of independent Poisson random variables is itself Poisson distributed, the total test count at time $t$ follows a Poisson distribution with mean $\lambda_{\text{Tot(t)}}$. Detection at time $t$ occurs when the observed count exceeds the baseline detection threshold, yielding:

$$\mathbb{P}_{Det}(t) = 1 - F_{\text{Pois}}\big(\mathfrak{T}; \lambda_{\text{Tot}}(t)\big) \quad \text{(Eq. B3)}$$

where $F_{Pois}$ denotes the Poisson cumulative distribution function (CDF) evaluated at the baseline threshold.

**B3. Cumulative and first-detection probabilities**

Assuming conditional independence across time periods given $\rho$ and $\tau$ (justified since the epidemic trajectory is treated as deterministic), the cumulative detection probability by time $t$ is:

$$F_{Det}(t \mid \rho, t > \tau) = 1 - s = \prod_{s=\tau}^{t}[1 - \mathbb{P}_{Det}(s)] \quad \text{(Eq. B4)}$$

The probability of first detection at exactly time $t$ is:

$$\mathbb{P}_{Det_1}(t) = \left(\prod_{s=\tau}^{t-1}[1 - \mathbb{P}_{Det}(s)]\right)\mathbb{P}_{Det}(t) \quad \text{(Eq. B5)}$$

#### B4. Expected detection time and median detection time

The *expected detection time* is computed as:

$$E[t_d] = \sum_t t \cdot P_{Det_1}(t) \quad \text{(Eq. B6)}$$

The median detection time ($t_{50}$) is defined as the first time at which the cumulative detection probability exceeds 0.5 ($F_{Det}(t|\rho, t > \tau) > 0.5$).

#### B5. Cumulative epidemic case estimates

Cumulative cases from epidemic occurrence at τ to a potential detection time $t$ are calculated by integrating the incidence curve (*I(u)*):

$$C(t) = \int_\tau^t I(u)\,du = \int_\tau^t exp(\rho(u-\tau)\,du = \frac{1}{\rho}(exp(\rho(t-\tau)-1) \quad \text{(Eq. B7)}$$

We subsequently compute the *expected cumulative case count* at detection as the sum of *C(t)* weighted by the probability of first detection at exactly time $t$ ($\mathbb{P}_{Det_1}(t)$):

$$\mathbb{E}[C(t_d)] = \sum_{t=\tau}^{\infty} C(t) \cdot P_{Det_1}(t) = \sum_{t=\tau}^{\infty} \underbrace{\frac{1}{\rho}(exp(\rho(t-\tau)-1)}_{C(t)} \times \underbrace{P_{Det_1}(t)}_{Probability\ of\ first\ detection\ at\ t \geq \tau} \quad \text{(Eq. B8)}$$

where the upper limit is computationally simulated as a finite time horizon *T*, chosen so that tail mass beyond *T* is negligible.

#### B6. Relationship to the general framework presented in the main text

The Poisson results serve as a lower bound on detection times: because the variance of a Poisson distribution is equivalent to its mean (φ = 1), the Poisson-based thresholds in framework are the tightest and detection is the most favorable. Introducing overdispersion (φ > 1) increases variance, raises detection thresholds, and delays detection. The general NB framework in the main text reduces exactly to these expressions when φ_Pos = φ_Neg = 1.

### Supplement C: Analytic Detection Framework: Convolution Approach for Negative Binomial Baselines

#### C1: Motivation and distributional assumptions

The proposed detection framework requires computing the probability that total observed test counts exceed a baseline threshold at each time after emergence of the novel epidemic. Under Poisson assumptions (Supplement B), the sum of independent Poisson random variables is itself Poisson — the total test count at any time $t$ follows a Poisson distribution whose mean is the sum of the component means.

This property does not hold for the negative binomial (NB) distribution: the sum of independent NB random variables with *different* dispersion parameters does not in general follow a known parametric distribution. In our framework, three independent components contribute to the total test count at time $t \geq \tau$ (post epidemic emergence):

$$X_{\text{Tot}}(t) = X_{\text{Pos}} + X_{\text{Neg}} + X_{\text{Epi}}(t)$$

where

$$X_{\text{Pos}} \sim \text{NB}(\mu_{\text{Pos}}, \varphi_{\text{Pos}})$$

$$X_{\text{Neg}} \sim \text{NB}(\mu_{\text{Neg}}, \varphi_{\text{Neg}})$$

$$X_{\text{Epi}}(t) \sim \text{Poisson}(\pi \cdot \exp(\rho[t - \tau]))$$

Here we parameterize the NB by its mean $\mu$ and variance-to-mean ratio (VMR) $\varphi$, so that $\text{Var}(X) = \mu\varphi$ and the standard NB size (dispersion) parameter is $r = \mu^2/(\mu\varphi - \mu) = \mu/(\varphi - 1)$ for $\varphi > 1$. When $\varphi_{\text{Pos}} \neq \varphi_{\text{Neg}}$ or when a Poisson component is added, the resulting sum does not follow a closed-form distribution but can be computed via numerical convolution.

*Note*: When $\varphi_{\text{Pos}} = \varphi_{\text{Neg}} = 1$, all three components are Poisson distributed and their sum is a Poisson distribution with rate parameter $\mu_{\text{Pos}} + \mu_{\text{Neg}} + \pi\exp(\rho[t - \tau])$, recovering the closed-form framework in Supplement B.

#### C2. Probability mass functions of individual test count components

For each test count component, we define the probability mass function (PMF) over non-negative integers $k = 0,1,2, \ldots$

##### Negative binomial components

Using the mean-VMR parameterization with size parameter $r = \mu/(\varphi - 1)$ and success probability $p = r/(r + \mu) = 1/\varphi$, the PMF of a negative binomial distribution is:

$$f_{\text{NB}}(k; \mu, \varphi) = \binom{k + r - 1}{k} \left(\frac{1}{\varphi}\right)^r \left(1 - \frac{1}{\varphi}\right)^k, \quad k = 0,1,2, \ldots$$

where $r = \mu/(\varphi - 1)$. For $\varphi = 1$, this reduces to a Poisson PMF with rate parameter $\mu$ (in the limiting case as $r \to \infty$).

##### Poisson epidemic component

The contribution of novel epidemic cases to total counts is assumed to follow a Poisson PMF:

$$f_{\text{Pois}}(k; \lambda(t)) = \frac{\lambda(t)^k e^{-\lambda(t)}}{k!}, \quad k = 0,1,2, \ldots$$

where $\lambda(t) = \pi \cdot \exp(\rho[t - \tau])$ is the time-varying expected number of epidemic-driven tests.

#### C3. Sequential convolution procedure for component PMFs

The PMF of the total test count $X_{\text{Tot}}(t)$ is obtained by sequentially convolving the PMFs of the independent test components. For two independent discrete random variables $U$ and $V$ with PMFs $f_U$ and $f_V$, the PMF of their sum $W = U + V$ can be computed as:

$$(f_U * f_V)(k) = \sum_{j=0}^{k} f_U(j) \cdot f_V(k - j), \quad k = 0,1,2, \ldots$$

We compute the composite PMF in two sequential steps:

**Step 1.** Convolve the two baseline components to obtain the PMF of the total baseline test count:

$$f_{\text{Base}}(k) = (f_{\text{Pos}} * f_{\text{Neg}})(k) = \sum_{j=0}^{k} f_{\text{NB}}(j; \mu_{\text{Pos}}, \varphi_{\text{Pos}}) \cdot f_{\text{NB}}(k - j; \mu_{\text{Neg}}, \varphi_{\text{Neg}})$$

This baseline PMF is assumed to be time-invariant and only needs to be computed once for a given parameter set.

**Step 2.** At each time post-emergence $t \geq \tau$, we convolve the baseline PMF with the time-varying epidemic contribution following a Poisson PMF to obtain the composite distribution:

$$f_{\text{Tot}}(k; t) = (f_{\text{Base}} * f_{\text{Epi},t})(k) = \sum_{j=0}^{k} f_{\text{Base}}(j) \cdot f_{\text{Pois}}(k - j; \lambda(t))$$

The resulting $f_{\text{Tot}}(k; t)$ is the exact PMF of total test counts at time $t$, from which all detection metrics are derived.

*Remark on negative test thresholds.* For analyses based solely on negative test counts, the convolution simplifies: the composite distribution at time $t$ is the convolution of the negative baseline NB component with the Poisson epidemic contribution component:

$$f_{\text{Neg,Tot}}(k; t) = (f_{\text{Neg}} * f_{\text{Epi},t})(k) = \sum_{j=0}^{k} f_{\text{NB}}(j; \mu_{\text{Neg}}, \varphi_{\text{Neg}}) \cdot f_{\text{Pois}}(k - j; \lambda(t))$$

Since positive baseline tests do not contribute to the negative-test detection threshold, Step 1 is not required, and the resulting two-component convolution is computationally less expensive.

#### C4: Computational implementation

Computationally, the PMFs are evaluated over a finite support $\{0,1, \ldots, K_{\max}\}$ where $K_{\max}$ is chosen so that the cumulative probability $\sum_{k=0}^{K_{\max}} f(k)$ exceeds $1 - \epsilon$ for a small tolerance (we used $\epsilon = 10^{-10}$). The convolution is then a finite sum and can be computed efficiently as:

1. **Pre-compute the baseline PMF vectors** $\mathbf{f}_{\text{Pos}} = [f_{\text{NB}}(0; \mu_{\text{Pos}}, \varphi_{\text{Pos}}), \dots, f_{\text{NB}}(K_{\text{Pos}}; \mu_{\text{Pos}}, \varphi_{\text{Pos}})]$ and $\mathbf{f}_{\text{Neg}}$ analogously.

2. **Convolve the baseline vectors** via the discrete convolution $\mathbf{f}_{\text{Base}} = \mathbf{f}_{\text{Pos}} * \mathbf{f}_{\text{Neg}}$, yielding a vector of length $K_{\text{Pos}} + K_{\text{Neg}} + 1$. This is computed once per parameter set.

3. **At each post-emergence time** $t \geq \tau$: Compute the Poisson PMF vector $\mathbf{f}_{\text{Epi},t}$ for the current epidemic mean $\lambda(t)$, then convolve: $\mathbf{f}_{\text{Tot},t} = \mathbf{f}_{\text{Base}} * \mathbf{f}_{\text{Epi},t}$.

4. **Estimate the detection probability:** The per-period detection probability at time $t$ is the upper tail of the composite distribution evaluated at the baseline detection threshold $\mathfrak{T}$:

$$\mathbb{P}_{\text{Det}}(t) = 1 - \sum_{k=0}^{\lfloor \mathfrak{T} \rfloor} f_{\text{Tot}}\,(k; t) = \sum_{k=\lfloor \mathfrak{T} \rfloor + 1}^{K_{\max}} f_{\text{Tot}}\,(k; t)$$

This is the exact NB counterpart of the detection probability based on the Poisson CDF in Supplement B (Eq. B3), replacing the closed-form Poisson CDF with the numerically computed complementary CDF of the convolved distribution.

All convolutions were implemented in R using vectorized operations. For the multidimensional parameter grid analyses, computations were parallelized across parameter combinations.

#### C5. Detection metrics based on convolved distributions

Given $\mathbb{P}_{\text{Det}}(t)$ from the convolved distributions, all subsequent detection metrics follow the same definitions as the Poisson case (Supplement B, Eqs. B4–B8), with no additional distributional assumptions beyond the per-period detection probabilities:

**Cumulative detection probability:**

$$F_{\text{Det}}(t \mid \rho, t > \tau) = 1 - \prod_{s=\tau}^{t} [\,1 - \mathbb{P}_{\text{Det}}(s)]$$

**First-detection probability:**

$$\mathbb{P}_{\text{Det}_1}(t) = \left( \prod_{s=\tau}^{t-1} [\,1 - \mathbb{P}_{\text{Det}}(s)] \right) \mathbb{P}_{\text{Det}}(t)$$

**Expected detection time:**

$$\mathbb{E}[t_d] = \sum_{t=\tau}^{T} t \cdot \mathbb{P}_{\text{Det}_1}(t)$$

**Expected cumulative cases at detection:**

$$\mathbb{E}[C(t_d)] = \sum_{t=\tau}^{T} C\,(t) \cdot \mathbb{P}_{\text{Det}_1}(t)$$

where $C(t) = \frac{1}{\rho}(\exp(\rho(t-\tau)) - 1)$ is the cumulative epidemic incidence at time $t$.

**Predictive intervals** (95% PI) are obtained as the 2.5th, 50th, and 97.5th percentiles (weighted quantiles) of the first-detection probability distribution $\mathbb{P}_{\text{Det}_1}(t)$, normalized over $t \geq \tau$.

**Median detection time** ($t_{50}$) is the earliest time at which $F_{\text{Det}}(t) > 0.5$.

*Note*: These expressions are distribution-agnostic with regards to the individual test count components — they depend on the per-period detection probabilities $\mathbb{P}_{\text{Det}}(t)$, which are derived from the convolved composite distributions. This modular structure allows the detection metric formulas to be identical under Poisson, NB, or any other distributional assumption for baseline counts; only the computation of $\mathbb{P}_{\text{Det}}(t)$ differs.

#### C6. Detection thresholds under negative binomial assumptions

The baseline detection threshold under the general NB framework is:

$$\mathfrak{T} = \mu_{\text{Base}} + \kappa \cdot \sigma_{\text{Base}}$$

**For total test thresholds:**

$$\mu_{\text{Base}} = \mu_{\text{Pos}} + \mu_{\text{Neg}}$$

$$\sigma^2_{\text{Base}} = \text{Var}(X_{\text{Pos}}) + \text{Var}(X_{\text{Neg}}) = \mu_{\text{Pos}}\varphi_{\text{Pos}} + \mu_{\text{Neg}}\varphi_{\text{Neg}}$$

$$\sigma_{\text{Base}} = \sqrt{\mu_{\text{Pos}}\varphi_{\text{Pos}} + \mu_{\text{Neg}}\varphi_{\text{Neg}}}$$

Note that the variance of the sum ($\sigma^2_{\text{Base}}$) equals the sum of variances by independence, even though the *distribution* of the sum is not NB. The threshold is computed from moments (mean and standard deviation), which are additive (assuming independence), not from the distributional form.

**For negative test thresholds:**

$$\mathfrak{T}_{\text{Neg}} = \mu_{\text{Neg}} + \kappa \cdot \sqrt{\mu_{\text{Neg}}\varphi_{\text{Neg}}}$$

**Poisson reference case** ($\varphi_{\text{Pos}} = \varphi_{\text{Neg}} = 1$):

$$\sigma_{\text{Base}} = \sqrt{\mu_{\text{Pos}} + \mu_{\text{Neg}}} = \sqrt{\mu_{\text{Base}}}$$

recovering the Poisson threshold $\mathfrak{T} = \mu_{\text{Base}} + \kappa\sqrt{\mu_{\text{Base}}}$ (cf. Supplement B, Eq. B1).
*Note* that mean := λ for Poisson per standard mathematical notation.

#### C7. Overview of the relationship between analytic and simulation approaches

The convolution-based analytic approach and the stochastic simulation framework (Supplement F) target the same quantities but differ in method:

| Quantity/Aspect | Analytic (convolution) | Simulation (Monte Carlo) |
|---|---|---|
| Distribution of total tests | Exact (numerically computed PMF) | Approximated from $s =$ 10,000 random draws |
| Detection probabilities | Exact tail probabilities from convolved PMF | Empirical frequencies of threshold exceedance |
| Detection time distribution | Exact first-detection probabilities | Empirical distribution of first threshold crossings |

| Quantity/Aspect | Analytic (convolution) | Simulation (Monte Carlo) |
|---|---|---|
| Computational cost | Scales with support size $K_{\max}$ and number of time points | Scales with number of simulations $s$ and parameter combinations |
| Uncertainty quantification | Derived from exact distributions | Includes Monte Carlo sampling variability |

The close agreement between analytic and simulation results (Tables 3–4 vs. Tables S1–S2) cross-validates both approaches. The analytic convolution approach provides exact results (up to numerical precision) and is preferred for systematic parameter exploration; the simulations serve as an independent corroboration and additionally yield empirical false positive rates (FP; 1 – specificity).

### D: Detailed Methods for Estimating the Underlying Epidemic Size from Routine Testing Data

#### D1. Mathematical framework

The framework estimates the size of the underlying emerging epidemic $N_{Epi}$ from observed test counts $X_{Obs}$, assumed to arise from two primary sources: routine baseline test counts with known mean $\mu_{base}$ and tests contributed by cases of the underlying epidemic. Given a testing fraction π among epidemic cases, the expected total test count is:

$$\mu_{total} = \mu_{base} + \pi \times N_{Epi}$$

#### D2. Analytic approach based on identity of Poisson and Chi-squared cumulative distribution functions

Assuming that test counts arise from a Poisson process, bounds and confidence intervals can be derived analytically via closed-form solutions based on the identity between Poisson and Chi-squared cumulative distribution functions. For a Poisson random variable *X* with rate (mean) parameter λ:

$$F_{Pois}(k;\lambda) = 1 - F_{\chi^2}\big(2\lambda; 2(k+1)\big) \quad for\ integer\ k, and$$

$$P(X = k) = F_{\chi^2}\big(2\lambda; 2(k+1)\big) - F_{\chi^2}(2\lambda; 2k)$$

From which we derive:

$$\mathrm{P(X \le k) = P\big(\chi^2_{2(k+1)} \ge 2\lambda\big)}$$

- where $\chi^2_{2(k+1)}$ denotes a Chi-squared distribution with df = 2 (k+1) degrees of freedom.

#### D2.1 Analytic exclusion bounds

Given a baseline detection threshold $\mathfrak{T} = \lambda_{base} + \kappa \times \sqrt{\lambda_{base}}$, where κ is the detection threshold sensitivity parameter, the maximum epidemic size consistent with not triggering an alert (at a pre-specified confidence level α is derived by:

1. Finding $\lambda_{total^{max}}$ such that $P(X \le \mathfrak{T}) = \alpha \quad (e.g., \alpha = 0.05)$
2. Using the Poisson-Chi-squared identity and setting: $\lambda_{total^{max}} = \frac{\chi^2_{2(\mathfrak{T}+1)}(1-\alpha)}{2}$
- Where $\chi^2_{2(\mathfrak{T}+1)}(1-\alpha)$ is derived from the Chi-squared quantile function
3. Converting the identified $\lambda_{total^{max}}$ to the upper bound of the epidemic size:

$$N_{Epi} = max\left(0, \frac{\lambda_{total^{max}} - \lambda_{base}}{\pi}\right)$$

This threshold-based exclusion bound addresses the question: "Given that there was no alert detected at this threshold, what underlying epidemic size can be excluded with $(1-\alpha)$ confidence?"

*Observation-based exclusion bounds*

For a specific observed test count $X_{Obs}$, the one-sided upper bound for the underlying epidemic size can be derived analogously from the Poisson-Chi-squared identity as:

1. $\lambda_{total^{upper}} = \frac{\chi^2_{2(X_{Obs}+1)}(1-\alpha)}{2}$ via the Chi-squared quantile function
2. $\lambda_{Epi^{upper}} = \lambda_{total^{upper}} - \lambda_{base}$

3. $N_{Epi^{upper}} = \frac{max\left(0, \lambda_{epidemic^{upper}}\right)}{\pi} = max\left(0, \frac{\lambda_{Epi^{upper}}}{\pi}\right)$

#### D2.2 Analytic two-sided confidence intervals

The $(1 - \alpha)$ confidence intervals are estimated based on the Poisson-Chi-squared identity as:

**Lower bound:**

$$\lambda_{total^{low}} = \frac{\chi^2_{2(X_{Obs}+1)}(\alpha/2)}{2}$$

Converted to the epidemic size bound: $N_{Epi^{lCI}} = max\left(0, \frac{\lambda_{total^{low}} - \lambda_{base}}{\pi}\right)$

**Upper bound:**

$$\lambda_{total^{up}} = \frac{\chi^2_{2(X_{Obs}+1)}(1 - \alpha/2)}{2}$$

Converted to the epidemic size bound: $N_{Epi^{uCI}} = max\left(0, \frac{\lambda_{total^{up}} - \lambda_{base}}{\pi}\right)$

*Note*: All estimated epidemic size bounds are truncated at zero to avoid implausible negative-valued epidemic sizes.

### D3. Simulation-based framework

#### D3.1 Stochastic algorithm

To account for overdispersion (variance > mean) commonly observed in real-world testing data, the baseline test counts are simulated using negative binomial distributions:

Positive tests: $NB\left(\mu_{pos}, \sigma^2_{pos}\right)$ with size (overdispersion) parameter $r_{pos} = \frac{\mu^2_{pos}}{\sigma^2_{pos} - \mu_{pos}}$

Negative tests: $NB\left(\mu_{neg}, \sigma^2_{neg}\right)$ with size (overdispersion) parameter $r_{neg} = \frac{\mu^2_{neg}}{\sigma^2_{neg} - \mu_{neg}}$

Note: the variance parameters $(\sigma^2_{pos}, \sigma^2_{neg})$ are derived from pre-specified variance-to-mean-ratio parameters $\varphi$:

$$\varphi = \frac{\sigma^2}{\mu} \Leftrightarrow \sigma^2 = \mu \times \varphi$$

The contribution of the underlying epidemic to the total test count is modeled based on a Poisson distribution for the expected test count arising from the epidemic:

Epidemic contribution $\sim Poisson\left(\pi \times N_{Epi}\right)$

This distributional assumption i) maps to the exponential epidemic growth model in the approaches previously employed to model this detection system (see Methods) and ii) follows from the approximation of the Poisson distribution to the Binomial distribution for count processes of rare events (under the Poisson Limit Theorem).

#### D3.2 Dynamic grid search

To determine the range of hypothetical sizes of the underlying epidemic, the simulation algorithm employs a dynamic grid search based on the simulated data for observed test counts ($X_{Obs}$):

1. The contribution of the underlying epidemic is estimated as: $\hat{\lambda}_{Epi} = max(0, X_{Obs} - \mu_{base})$
2. From which the expected epidemic size is calculated: $\mathbb{E}[N_{Epi}] = \hat{\lambda}_{Epi}/\pi$
3. The maximum of the grid is set to:

$$Grid_{max} = max\left(3000, \mathbb{E}[N_{Epi}] \times b, \frac{X_{Obs} - \mu_{base} + 3 \times \sigma_{base}}{\pi}\right)$$

Where $b$ is a pre-specified buffer ($b \geq 2$) to ensure adequate coverage of the search grid, and $\sigma_{base} = \sqrt{(\sigma^2{}_{pos} + \sigma^2{}_{neg})}$. The grid maximum is set to a minimum of 3,000 to avoid missing epidemic sizes for extreme scenarios (e.g., very low $\pi$).

#### D3.3 Epidemic size estimation and likelihood profile

*Closed-form maximum likelihood estimator*

Under our model (cf. Methods), the maximum likelihood estimate (MLE) has a closed-form solution:

$$\hat{N}_{Epi_{MLE}} = max(0, (X_{Obs} - \mu_{base})/\pi)$$

where $X_{Obs}$ denotes the observed test count, $\mu_{base} = \mu_{pos} + \mu_{neg}$ is the expected total baseline test count, and $\pi$ is the probability of testing among epidemic cases. This analytic estimator is unbiased and independent of the baseline variance structure (i.e., identical under Poisson and negative binomial assumptions). The MLE derived from the simulated likelihood profiles (see below) targets the same parameter and is equivalent in expectation, so we chose the closed-form estimator for our main results (Figure 3 and 4).

*Simulation-based likelihood profile*

To properly account for baseline variability and overdispersion, uncertainty was quantified via simulated likelihood profiles. For each hypothetical epidemic size $N_{Epi[i]}$ in the search grid, the simulation algorithm:

1. Generates $s$ = 10,000 realizations of total test counts ($X_s$) by combining random draws from a negative binomial baseline with Poisson epidemic contributions:

$$X_s = X_{pos,s} + X_{neg,s} + X_{Epi,s} where$$

$$X_{pos,s} \sim NB(\mu_{pos}, \sigma^2_{pos})$$

$$X_{neg,s} \sim NB(\mu_{neg}, \sigma^2_{neg})$$

$$X_{Epi,s} \sim Poisson(\pi \times N_{Epi[i]})$$

2. Estimates the likelihood of observing an epidemic size ($N_{Epi[i]}$) given the observed test count ($X_{Obs}$) using a rectangular kernel:

$$\mathcal{L}\left(N_{Epi[i]} \middle| X_{Obs}\right) = \frac{1}{s} \times \sum_{s} \frac{\mathbb{I}(|X_s - X_{Obs}| \leq w)}{2w + 1}$$

where $w = max(1, \sigma_{base}/5)$ is the width of the kernel window and $\mathbb{I}(\cdot)$ is the indicator function.

The resulting likelihood profile $\mathcal{L}\left(N_{Epi} \middle| X_{Obs}\right)$ across the grid of epidemic sizes can be used to derive the maximum likelihood estimate of the epidemic size ($\widehat{N}_{Epi_{MLE}} = arg \max_{N_{Epi[i]}} \mathcal{L}\left(N_{Epi[i]} \middle| X_{Obs}\right)$) and provides the foundation for all uncertainty quantification procedures described in D3.4.

*Normalized likelihood distribution*

A normalized probability distribution can be obtained from the likelihood profile as:

$$\mathbb{P}\left(N_{Epi[i]} \middle| X_{Obs}\right) = \frac{\mathcal{L}\left(N_{Epi[i]} \middle| X_{Obs}\right)}{\sum_j \mathcal{L}\left(N_j \middle| X_{Obs}\right)}$$

where $N_{Epi[i]}$ represents a specific epidemic size being evaluated (e.g., $N_{Epi} = 300$) and $N_j$ represents the vector of all epidemic sizes in the search grid.

#### D3.4 Uncertainty quantification

**Test-inversion confidence intervals**

We derive 95% confidence intervals for the underlying epidemic size ($N_{Epi}$) using a test-inversion approach. For each hypothesized epidemic size $N_{Epi}$ in the evaluated search grid, we test the null hypothesis $H_0$: $N_{Epi} = N$ by computing the probability of observing a test count as or more extreme than $X_{Obs}$, and generate the Monte Carlo tail probabilities of the sampling distribution of total tests ($X$) under the $H_0$:

$$p_L(N) = P\left( X \leq X_{Obs} \mid N = N_{Epi} \right); p_R(N) = P\left( X \geq Xobs \mid N = N_{Epi} \right)$$

From these, we form a two-sided p-value:

$$p_2(N) = 2 \times min\{p_L(N), p_R(N)\}$$

The (1-α) confidence interval includes all $N$ values where $p_2(N) \geq \alpha$:

$$CI_{1-\alpha} = \{N: p_2(N) \geq \alpha\}$$

with endpoints determined by linear interpolation between adjacent grid points where the p-value crosses the significance threshold (α = 0.05 for 95%CI).

**Bootstrap confidence interval for epidemic size based on likelihood profile:**

To validate the test-inversion CI, we also obtain 95%CI via parametric bootstrap using the closed-form MLE ($\widehat{N}_{Epi_{MLE}}$) as the fitted value:

1. Under the fitted model with $N_{Epi} = \widehat{N}_{Epi}$, we generate bootstrap totals:
   $X^{*(b)} = X_{\text{pos}}^{*(b)} + X_{\text{neg}}^{*(b)} + X_{\text{epi}}^{*(b)}$,
   where $X_{\text{pos}}^{*(b)} \sim \text{NB}(\mu_{\text{pos}}, \text{var}_{\text{pos}}), X_{\text{neg}}^{*(b)} \sim \text{NB}(\mu_{\text{neg}}, \text{var}_{\text{neg}}), X_{\text{epi}}^{*(b)} \sim \text{Poisson}(\pi \times \widehat{N})$;
2. For each bootstrap replicate $b = 1, \dots, B$, we re-estimate $\widehat{N}_{Epi}{}^{*(b)}$ using the same closed-form MLE applied to $X^{*(b)}$;
3. With $\alpha = 0.05$, we estimate the 95% confidence intervals based on quantiles ($Q$):

$$\text{CI}_{0.95}(N_{Epi}) = [\, Q_{\alpha/2}\{\widehat{N}_{Epi}{}^{*(b)}\}, Q_{1-\alpha/2}\{\widehat{N}_{Epi}{}^{*(b)}\}\,]$$

   where the lower bound $Q_{\alpha/2}\{\widehat{N}_{Epi}{}^{*(b)}\}$ is truncated at 0 (to avoid implausible values).

**Range of compatible epidemic size (based on Likelihood Ratio Test)**

Following Wilks' theorem, the log-likelihood ratio statistic (LR) asymptotically approaches a Chi-squared distribution under the null (as the sample size approaches ∞): $-2log(LR) \sim \chi_1^2$

The set of epidemic sizes compatible with the observed test count can thus be obtained from the likelihood profile as:

$$Compatible\ set\ = \left\{ \frac{N_{Epi[i]} : \mathcal{L}\left(N_{Epi[i]}\right)}{\mathcal{L}\left(N_{Epi_{MLE}}\right)} \geq exp\left(\frac{-\chi_1^2(0.95)}{2}\right) \right\}$$

where $\chi_1^2(0.95) = 3.841$, yielding a LR threshold ≈ 0.147.

### E: Empirical Assessment of Overdispersion in Surveillance Data

To contextualize the variance-to-mean ratio range ($\varphi \in [1,5]$) explored in the main analyses (cf. Tables 2–4 and Supplements B–C), we empirically characterized the overdispersion of routine respiratory virus testing data published by the U.S. Centers for Disease Control and Prevention (CDC) National Respiratory and Enteric Virus Surveillance System (NREVSS). NREVSS is a voluntary laboratory-based system in which approximately 300 participating U.S. clinical and public health laboratories report weekly aggregate counts of tests performed and pathogen-specific positive detections. These reports map directly onto the data structure assumed in our framework—total tests partitioned into positive and negative components—making them an ideal empirical reference for calibrating $\varphi_{\text{Pos}}$ and $\varphi_{\text{Neg}}$.

#### E1. Data sources and pre-processing

Weekly counts were obtained via the data.cdc.gov SODA API for two respiratory pathogens with continuous reporting through the analytic window:

- **SARS-CoV-2** nucleic acid amplification tests (NAATs; March 2020 – March 2026).
- **Respiratory syncytial virus (RSV)** PCR tests (April 2020 – March 2026).

Records were extracted at both the national level and for all ten U.S. Department of Health and Human Services (HHS) regions; results presented below correspond to the national series and to three representative regions (HHS Regions 1, 4, and 9; results for the remaining regions were concordant). For each pathogen, weekly negative-test counts were derived as $X_{\text{Neg},t} = X_{\text{Tot},t} - X_{\text{Pos},t}$.

*Note on data cleaning and transformation (RSV total-test back-calculation).* In the public NREVSS PCR release for RSV, the field reporting total PCR tests performed (`pcr_tests`) was null for all observations. We therefore back-calculated the total weekly test volume from the two reported fields—PCR detections and percent positivity—using (rounded to the nearest integer):

$$X_{\text{Tot},t}^{(\text{RSV})} = \left\lfloor \frac{X_{\text{Pos},t}^{(\text{RSV})}}{p_t/100} \right\rceil, \quad p_t > 0 \quad (\text{Eq. E1})$$

where $p_t$ is the reported weekly test positivity (in percent). Records with $p_t = 0$ (no detection) cannot be back-solved and were excluded from the variance computations. After data cleaning, the deduplicated SARS-CoV-2 series comprised $N = 314$ national weeks and the RSV series $N = 304$ national weeks.

#### E2. Definition of stable baseline periods

Because our framework uses the *baseline* (pre-emergence) variance of routine testing data, we restricted analyses to weeks plausibly representing endemic (i.e., without major epidemics) conditions for each pathogen:

- **SARS-CoV-2 stable period:** weeks with test positivity below 5% *and* posted on or after 1 January 2022 (i.e., after the initial Omicron wave). $n = 77$ weeks on the national level.
- **RSV stable period:** weeks falling in the off-season months of May through September *and* posted from 2021 onward, excluding the post-pandemic rebound transition period. $n = 109$ weeks on the national level.

These criteria mirror common practice in surveillance baseline estimation (low-positivity windows for endemic respiratory pathogens; off-season months for seasonal pathogens) and were applied identically at the national and regional levels.

#### E3. Overdispersion metrics

Three complementary statistics were computed for the negative-test series at each aggregation level, with results for total tests verified in parallel to yield similar conclusions.

*E3.1 Raw variance-to-mean ratio.* The unconditional, sample-based estimator of the VMR (φ) is:

$$\hat{\varphi}_{\text{raw}} = \frac{\widehat{\text{Var}}(X_t)}{\widehat{\mathbb{E}}(X_t)} \quad \text{(Eq. E2)}$$

computed across all weeks within each stable period. This estimator captures the total dispersion of the count series, including any systematic temporal trend or level shifts that survived the stable-period filter.

*E3.2 Detrended VMR and coefficient of variation.* To isolate stochastic, week-to-week variability from residual seasonality and secular trends in reporting volume, we detrended time series using a 4-week centered moving average ($\bar{X}_t^{(4)}$) and recomputed the VMR on the residuals while normalizing by the original-series mean (to preserve interpretability on the count scale):

$$\bar{X}_t^{(4)} = 1/4 \sum_j X_{t+j}, \quad \varepsilon_t = X_t - \bar{X}_t^{(4)} \quad \text{(Eq. E3)}$$

$$\hat{\varphi}_{\text{det}} = \frac{\widehat{\text{Var}}(\varepsilon_t)}{\widehat{\mathbb{E}}(X_t)}, \qquad \widehat{CV}_{\text{det}} = \frac{\hat{\sigma}(\varepsilon_t)}{\widehat{\mathbb{E}}(X_t)} \quad \text{(Eq. E4)}$$

The resulting detrended coefficient of variation (CV) provides a scale-free measure of week-to-week stochastic noise that is approximately invariant to the aggregation level of the time series.

*E3.3 Estimated site-level $\varphi$ via scale projection.* Because public NREVSS releases do not disaggregate below the HHS regional level, we cannot directly observe $\varphi$ at the facility or sentinel-site level at which our framework is likely to operate. We therefore projected $\varphi$ to hypothetical sites with mean weekly negative-test volume $\mu_{\text{site}}$ under the assumption that the stochastic CV is approximately scale-invariant across aggregation levels (a conservative assumption supported by the empirical observation that regional CVs were similar to or larger than national CVs; cf. Table E2). Under this assumption, $\sigma_{\text{site}} \approx CV \cdot \mu_{\text{site}}$, so $\sigma_{\text{site}}^2 \approx CV^2 \cdot \mu_{\text{site}}^2$, and consequently:

$$\varphi_{\text{site}} = \frac{\sigma_{\text{site}}^2}{\mu_{\text{site}}} \approx \mu_{\text{site}} \times CV_{\text{det}}^2 \quad \text{(Eq. E5)}$$

Equation E5 makes the scaling explicit: provided the relative noise level is approximately constant across aggregation scales, $\varphi$ scales in direct proportion to the mean (as a *linear* function of CV, compared to constant $\varphi = 1$ under Poisson assumptions). A given $CV_{\text{det}}$ thus implies a one-dimensional curve mapping site-level $\varphi$ values to site-level mean counts ($\mu_{\text{site}}$), against which the manuscript's modeled range can be benchmarked.

*E3.4 Week-to-week volatility.* As an additional model-free check, we computed the median absolute weekly percent change, $\mathrm{med}_t|\Delta X_t/X_{t-1}| \times 100\%$, and compared it against the Poisson reference value. For two independent Poisson observations with common mean $\mu$, the standard deviation of their difference is $\sqrt{2\mu}$, yielding an expected percent change on the order of $\sqrt{2/\mu} \times 100\%$. The ratio of observed to expected percent change provides a non-parametric estimate of excess week-to-week variability relative to a pure Poisson process.

### E4. Results

*E4.1 Raw and detrended VMR by aggregation level.* At the national scale during stable periods, raw VMRs for negative tests were several orders of magnitude above the range modeled in the manuscript ($\hat{\varphi}_{\mathrm{raw}} \approx 41{,}190$ for SARS-CoV-2 and $\hat{\varphi}_{\mathrm{raw}} \approx 3{,}885$ for RSV; Table E1). Removal of the 4-week moving average reduced these values by roughly an order of magnitude ($\hat{\varphi}_{\mathrm{det}} \approx 1{,}864$ and $\hat{\varphi}_{\mathrm{det}} \approx 230$, respectively), indicating that a substantial fraction of the raw variance reflects systematic level changes (laboratory participation drift, reporting policy shifts, residual seasonality) rather than week-to-week stochasticity. At the regional scale, both raw and detrended VMRs were substantially smaller (raw $\hat{\varphi} = 2{,}221$–$7{,}709$ and detrended $\hat{\varphi} = 91$–$195$ for SARS-CoV-2; raw $\hat{\varphi} = 510$–$1{,}323$ and detrended $\hat{\varphi} = 95$–$136$ for RSV), consistent with a strong scale dependence: as both $\mu$ and $\sigma$ contract under disaggregation, $\varphi = \sigma^2/\mu$ contracts approximately linearly with $\mu$ (Eq. E5).

The corresponding national detrended CVs were $CV_{\mathrm{det}} \approx 0.147$ for SARS-CoV-2 and $\approx 0.078$ for RSV. Regional detrended CVs were comparable to or higher than the national value (SARS-CoV-2: 0.11–0.17; RSV: 0.15–0.22), supporting the use of the national CV as a *conservative lower bound* for the projected site-level $\varphi$.

*E4.2 Implied site-level $\varphi$.* Applying Eq. E5 with the national detrended CVs across a grid of plausible site-level mean weekly volumes (Table E2) reproduced the regimes assumed in the main analyses. For SARS-CoV-2, the modeled range ($\varphi \in [1,5]$) corresponds to mean weekly negative-test volumes of $\mu_{\mathrm{site}} \approx 50$–$230$. For RSV, the same $\varphi$ range corresponds to $\mu_{\mathrm{site}} \approx 165$–$820$, reflecting its lower stochastic CV. At broader aggregation scales ($\mu \gtrsim 1{,}000$), implied $\varphi$ exceed the modeled range, indicating that systematic, non-stochastic variability (e.g., laboratories with different testing capacity joining or leaving the reporting network) dominates and that simple compartmental NB models are no longer adequate without additional trend correction (e.g., Farrington or state-space algorithms).

*E4.3 Week-to-week volatility relative to Poisson.* Median absolute weekly percent change in negative tests was $\approx$ 5.8% for SARS-CoV-2 and $\approx$ 6.9% for RSV at the national level, against Poisson references $< 1\%$ at those scales (excess factors $\approx 12\times$ and $\approx 9\times$, respectively). At the HHS regional level, observed percent changes were modestly larger (6–13%) but Poisson references were also larger due to the smaller mean counts, yielding excess factors of $\approx$ 3–5 $\times$. Even at the regional scale, the observed week-to-week variability is thus incompatible with a Poisson baseline, corroborating the convolution-based detection probabilities described in Supplement C.

**Table E1.** Empirical mean and overdispersion of weekly negative-test counts during stable periods, by pathogen and aggregation level.

| Level | SARS-CoV-2 mean ($\hat{\mu}$) | Raw VMR ($\hat{\varphi}_{\mathrm{raw}}$) | Detrended VMR ($\hat{\varphi}_{\mathrm{det}}$) | RSV mean ($\hat{\mu}$) | Raw VMR | Detrended VMR |
|---|---|---|---|---|---|---|
| National | 85,922 | 41,190 | 1,864 | 37,643 | 3,885 | 230 |

| | | | | | | |
|---|---|---|---|---|---|---|
| Region 1 | 6,702 | 5,921 | 165 | 2,282 | 510 | 106 |
| Region 4 | 7,207 | 2,221 | 91 | 4,320 | 720 | 95 |
| Region 9 | 6,946 | 7,709 | 195 | 3,788 | 1,323 | 136 |

*Note.* SARS-CoV-2 stable period: weeks with test positivity $< 5\%$ posted from 1 January 2022 onward. RSV stable period: May–September weeks posted from 2021 onward. Detrended VMR computed as $\widehat{\mathrm{Var}}(\varepsilon_t)/\widehat{\mathbb{E}}(X_t)$ with $\varepsilon_t$ defined in Eq. E3. Three HHS regions are shown as representative; the remaining seven regions yielded similar values.

**Table E2.** Implied site-level overdispersion ($\hat{\varphi}_{\text{site}} = \mu_{\text{site}} \times CV^2_{\text{det}}$, Eq. E5) as a function of mean weekly negative-test volume, computed using the national detrended CV.

| $\mu_{\text{site}}$ | SARS-CoV-2 $\hat{\varphi}_{\text{site}}$ | Classification of dispersion | RSV $\hat{\varphi}_{\text{site}}$ | Classification |
|---|---|---|---|---|
| 30 | 0.65 | ≈ Poisson* | 0.18 | ≈ Poisson* |
| 50 | 1.08 | ≈ Poisson* | 0.31 | ≈ Poisson* |
| 100 | 2.17 | Moderate overdispersion (2–5) | 0.61 | ≈ Poisson* |
| 200 | 4.34 | Moderate overdispersion (2–5) | 1.22 | Low overdispersion (1–2) |
| 500 | 10.9 | High overdispersion (> 5) | 3.06 | Moderate overdispersion (2–5) |
| 1,000 | 21.7 | High overdispersion (> 5) | 6.12 | High overdispersion (5–10) |

**Poisson models were used as the lower bound in the presented framework, so underdispersion was not explicitly modeled.*

*Note:* CV values: SARS-CoV-2 $CV_{\text{det}} = 0.147$; RSV $CV_{\text{det}} = 0.078$ (national, stable periods, Section E4.1). Classifications follow the manuscript's ranges: $\varphi \leq 1$, ≈ Poisson; $1 < \varphi \leq 2$, low overdispersion; $2 < \varphi \leq 5$, moderate overdispersion; $\varphi > 5$, high overdispersion. Regional detrended CVs were comparable or higher (Section E4.1), so these projections represent conservative lower bounds for site-level $\varphi$.

#### E5. Implications for the modeled $\varphi$ range and limitations

These empirical results support four conclusions that anchor the modeling choices presented in the main text. First, the modeled overdispersion range ($\varphi \in [1,5]$) is empirically realistic for individual reporting facilities and small sentinel networks operating at mean weekly negative-test volumes of approximately 50–500—precisely the regime in which the proposed early-detection framework is likely to be deployed. Second, at broader aggregation scales (regional, state, national), $\varphi$ rises rapidly with $\mu$ (Eq. E5), and the dominant variance source shifts from stochastic count noise to systematic shifts in laboratory participation (e.g., size and testing capacity) and testing policy; simplified NB compartmental models without additional trend correction are unlikely to adequately describe such large-scale aggregates. Third, the observed week-to-week volatility exceeds the Poisson reference by 3–12× at every aggregation level examined, confirming that the negative binomial extension developed in Supplement C is necessary for any realistic surveillance setting. Fourth, the demonstrated inverse coupling between aggregation scale and $\varphi$ implies that *spatial disaggregation of surveillance* is itself a variance-reducing intervention that moves the data into the favorable detection regime characterized in the main analyses.

A principal limitation of these exploratory analyses is that the site-level projections in Table E2 rely on the assumption that the stochastic CV is approximately invariant across levels of disaggregation. The empirical regional-versus-national comparison provides indirect evidence supporting this assumption, but direct estimation of $\varphi$ from facility-level NREVSS records—currently unavailable in the public release—would yield more definitive calibration. A second limitation is that the back-calculation of total RSV tests from reported detections and percent positivity (Eq. E1) inherits any rounding in the underlying CDC figures; sensitivity analyses excluding low-volume weeks ($p_t < 0.5\%$, where back-calculation is most sensitive) yielded VMR estimates within 5% of the values in Tables E1–E2.

**F: Stochastic Simulation Methods**

We developed a stochastic simulation framework to corroborate the analytic approaches under both negative binomial and Poisson assumptions and conducted simulations for total and negative test counts. Test counts were simulated over a specified horizon T (e.g., $t = 156$), with pre-epidemic baseline data (e.g., $t = 1–51$) generated via two independent negative binomial (or Poisson) processes for positive and negative tests. Baseline means and standard deviations were treated as known (i.e., deterministic).

At time $\tau$ (e.g., $\tau = 52$), an exponentially growing, deterministic epidemic of the novel disease is introduced, augmenting total test counts via a Poisson-distributed random variable with mean equal to the product of incident cases and probability $\pi$. An outbreak is detected when test counts exceed the baseline threshold; detections prior to $\tau$ are classified as false positives. Each set of parameters (Table 1) was simulated over *s = 10,000* iterations with detection metrics (e.g., median detection time, false positive rates, and expected cumulative cases at detection) aggregated across iterations to comparatively evaluate detection performance under different parameter values and distributional assumptions.

**Supplementary Tables**

**Table S1: Simulation Estimates of Outbreak Detection Performance Using Total Test Thresholds, including Poisson reference estimates**

| Parameter | Value | Distribution | Threshold | Median detection time ($t_{md}$) [a] | Median Cumulative Cases at detection | Mean detection time [a] [95%PI] | Mean Cumulative Cases at detection [95%PI] | FP [b] |
|---|---|---|---|---|---|---|---|---|
| $\mu_{Positive}$ | | | | | | | | |
| | 30 | NB | 172 | 29 | 1647 | 28 [9; 32] | 1633 [25; 3004] | 0.121 |
| | | Poisson | 165 | 28 | 1347 | 27 [13; 31] | 1398 [62; 2459] | 0.089 |
| | 100 (ref) | NB | 252 | 30 | 2012 | 29 [9; 33] | 2041 [25; 3671] | 0.107 |
| | | Poisson | 243 | 29 | 1647 | 28 [13; 32] | 1721 [62; 3004] | 0.085 |
| | 300 | NB | 474 | 32 | 3004 | 31 [12; 35] | 2991 [50; 5478] | 0.093 |
| | | Poisson | 460 | 31 | 2459 | 30 [13; 34] | 2432 [62; 4484] | 0.090 |
| | 1000 | NB | 1222 | 35 | 5478 | 33 [14; 37] | 4958 [77; 8175] | 0.088 |
| | | Poisson | 1200 | 34 | 4484 | 33 [16; 36] | 4122 [118; 6692] | 0.073 |
| $\mu_{Negative}$ | | | | | | | | |
| | 30 | NB | 172 | 29 | 1647 | 28 [8; 32] | 1616 [20; 3004] | 0.121 |
| | | Poisson | 165 | 28 | 1347 | 27 [12; 31] | 1395 [50; 2459] | 0.089 |
| | 100 (ref) | NB | 252 | 30 | 2012 | 29 [9; 33] | 2012 [25; 3671] | 0.107 |
| | | Poisson | 243 | 29 | 1647 | 28 [13; 32] | 1721 [62; 3004] | 0.085 |
| | 300 | NB | 474 | 32 | 3004 | 31 [13; 35] | 2969 [62; 5478] | 0.090 |
| | | Poisson | 460 | 31 | 2459 | 30 [13; 34] | 2425 [50; 4484] | 0.089 |
| | 1000 | NB | 1222 | 35 | 5478 | 33 [15; 37] | 4945 [95; 8175] | 0.087 |
| | | Poisson | 1200 | 34 | 4484 | 33 [16; 36] | 4151 [118; 6692] | 0.075 |
| $\varphi_{Positive}$ | | | | | | | | |
| | 1.5 (ref) | NB | 252 | 30 | 2012 | 29 [9; 33] | 2039 [25; 3671] | 0.107 |
| | 3 | NB | 264 | 31 | 2459 | 30 [8; 34] | 2488 [20; 4484] | 0.129 |
| | 5 | NB | 277 | 32 | 3004 | 30 [6; 35] | 2918 [12; 5478] | 0.166 |
| $\varphi_{Negative}$ | | | | | | | | |
| | 1.5 (ref) | NB | 252 | 30 | 2012 | 29 [9; 33] | 2039 [25; 3671] | 0.107 |

| | | | | | | | | |
|---|---|---|---|---|---|---|---|---|
| | 3 | NB | 264 | 31 | 2459 | 30 [9; 34] | 2487 [25; 4484] | 0.132 |
| | 5 | NB | 277 | 32 | 3004 | 30 [7; 35] | 2933 [15; 5478] | 0.170 |
| **π** | | | | | | | | |
| | 0.10 (ref) | NB | 252 | 30 | 2012 | 29 [9; 33] | 2040 [25; 3671] | 0.106 |
| | | Poisson | 243 | 29 | 1647 | 28 [13; 32] | 1721 [62; 3004] | 0.085 |
| | 0.15 | NB | 252 | 28 | 1347 | 27 [9; 31] | 1367 [25; 2459] | 0.106 |
| | | Poisson | 243 | 27 | 1102 | 27 [12; 30] | 1148 [50; 2012] | 0.085 |
| | 0.20 | NB | 252 | 27 | 1102 | 25 [9; 30] | 1022 [25; 2012] | 0.106 |
| | | Poisson | 243 | 26 | 901 | 25 [12; 29] | 866 [50; 1647] | 0.085 |
| **ρ** | | | | | | | | |
| | 0.15 | NB | 252 | 40 | 2683 | 37 [9; 44] | 2397 [19; 4894] | 0.106 |
| | | Poisson | 243 | 38 | 1986 | 36 [13; 42] | 2030 [40; 3624] | 0.085 |
| | 0.20 (ref) | NB | 252 | 30 | 2012 | 29 [9; 33] | 2040 [25; 3671] | 0.106 |
| | | Poisson | 243 | 29 | 1647 | 28 [13; 32] | 1721 [62; 3004] | 0.085 |
| | 0.225 | NB | 252 | 27 | 1928 | 26 [9; 30] | 1911 [29; 3791] | 0.106 |
| | | Poisson | 243 | 26 | 1539 | 25 [13; 29] | 1610 [78; 3027] | 0.085 |
| **κ** | | | | | | | | |
| | 2 | NB | 235 | 21 | 328 | 18 [0; 31] | 591 [0; 2459] | 0.737 |
| | | Poisson | 229 | 21 | 328 | 18 [1; 30] | 542 [1; 2012] | 0.703 |
| | 3 (ref) | NB | 252 | 30 | 2012 | 29 [9; 33] | 2040 [25; 3671] | 0.106 |
| | | Poisson | 243 | 29 | 1647 | 28 [13; 32] | 1721 [62; 3004] | 0.085 |
| | 4 | NB | 270 | 33 | 3671 | 32 [28; 35] | 3370 [1347; 5478] | 0.005 |
| | | Poisson | 257 | 32 | 3004 | 31 [28; 34] | 2754 [1347; 4484] | 0.003 |

*Abbreviations*: FP: false positive rate; NB: negative binomial; 95%PI: 95% empirical predictive intervals derived from Monte Carlo simulations to estimate the probability of first detection; ref: reference value; $t_{md}$: median detection time; $t_E$: expected time to detection

[a] Time units correspond to the surveillance system's reporting period (daily or weekly in most jurisdictions) and need to be interpreted in the context of the monitored pathogen. For systems with daily cadence monitoring rapidly propagating pathogens (e.g., daily growth rates of 0.20), this represents 28-35 days to detection. For weekly reporting systems monitoring slower-spreading pathogens, this could represent weeks.

[b] False positive rate computed based on *s = 10,000* simulations

Table S2: Simulation Estimates of Outbreak Detection Performance Using Negative Test Thresholds, including Poisson reference

| Parameter | Value | Distribution | Threshold | Median detection time ($t_{md}$) [a] | Median Cumulative Cases at detection | Mean detection time [a] [95%PI] | Mean Cumulative Cases at detection [95%PI] | FP [b] |
|---|---|---|---|---|---|---|---|---|
| $\mu_{Positive}$ | | | | | | | | |
| | 30 | NB | 137 | 28 | 1347 | 27 [9; 31] | 1436 [25; 2459] | 0.132 |
| | | Poisson | 130 | 27 | 1102 | 26 [10; 30] | 1160 [32; 2012] | 0.105 |
| | 100 (ref) | NB | 137 | 28 | 1347 | 27 [9; 31] | 1426 [25; 2459] | 0.129 |
| | | Poisson | 130 | 27 | 1102 | 26 [10; 30] | 1161 [32; 2012] | 0.107 |
| | 300 | NB | 137 | 28 | 1347 | 27 [9; 31] | 1428 [25; 2459] | 0.129 |
| | | Poisson | 130 | 27 | 1102 | 26 [10; 30] | 1153 [32; 2012] | 0.110 |
| | 1000 | NB | 137 | 28 | 1347 | 27 [9; 31] | 1424 [25; 2459] | 0.128 |
| | | Poisson | 130 | 27 | 1102 | 26 [11; 30] | 1154 [40; 2012] | 0.112 |
| $\mu_{Negative}$ | | | | | | | | |
| | 30 | NB | 51 | 25 | 737 | 24 [7; 29] | 787 [15; 1647] | 0.147 |
| | | Poisson | 47 | 24 | 603 | 23 [8; 28] | 634 [20; 1347] | 0.112 |
| | 100 (ref) | NB | 137 | 28 | 1347 | 27 [9; 31] | 1426 [25; 2459] | 0.129 |
| | | Poisson | 130 | 27 | 1102 | 26 [10; 30] | 1161 [32; 2012] | 0.107 |
| | 300 | NB | 364 | 31 | 2459 | 30 [12; 34] | 2556 [50; 4484] | 0.098 |
| | | Poisson | 352 | 30 | 2012 | 29 [13; 33] | 2093 [62; 3671] | 0.086 |
| | 1000 | NB | 1117 | 34 | 4484 | 33 [15; 37] | 4776 [95; 8175] | 0.082 |
| | | Poisson | 1095 | 33 | 3671 | 32 [16; 36] | 3908 [118; 6698] | 0.075 |
| $\varphi_{Positive}$ | | | | | | | | |
| | 1.5 (ref) | NB | 137 | 28 | 1347 | 27 [9; 31] | 1428 [25; 2459] | 0.127 |
| | 3 | NB | 137 | 28 | 1347 | 27 [10; 31] | 1426 [32; 2459] | 0.128 |
| | 5 | NB | 137 | 28 | 1347 | 27 [10; 31] | 1432 [32; 2459] | 0.128 |
| $\varphi_{Negative}$ | | | | | | | | |
| | 1.5 (ref) | NB | 137 | 28 | 1347 | 27 [9; 31] | 1428 [25; 2459] | 0.127 |
| | 3 | NB | 152 | 30 | 2012 | 28 [6; 33] | 1948 [12; 3671] | 0.180 |
| | 5 | NB | 168 | 31 | 2459 | 29 [5; 34] | 2529 [9; 4484] | 0.205 |
| $\pi$ | | | | | | | | |

| | | | | | | | | |
|---|---|---|---|---|---|---|---|---|
| | 0.10 (ref) | NB | 137 | 28 | 1347 | 27 [9; 31] | 1426 [25; 2459] | 0.129 |
| | | Poisson | 130 | 27 | 1102 | 26 [10; 30] | 1161 [32; 2012] | 0.107 |
| | 0.15 | NB | 137 | 26 | 901 | 25 [9; 29] | 959 [25; 1647] | 0.129 |
| | | Poisson | 130 | 25 | 737 | 24 [9; 28] | 779 [25; 1347] | 0.107 |
| | 0.20 | NB | 137 | 25 | 737 | 24 [9; 28] | 717 [25; 1347] | 0.130 |
| | | Poisson | 130 | 24 | 603 | 23 [9; 27] | 583 [25; 1102] | 0.107 |
| **ρ** | | | | | | | | |
| | 0.15 | NB | 137 | 37 | 1708 | 35 [9; 41] | 1664 [19; 3118] | 0.129 |
| | | Poisson | 130 | 36 | 1469 | 34 [11; 40] | 1360 [28; 2683] | 0.107 |
| | 0.20 (ref) | NB | 137 | 28 | 1347 | 27 [9; 31] | 1426 [25; 2459] | 0.130 |
| | | Poisson | 130 | 27 | 1102 | 26 [10; 30] | 1161 [32; 2012] | 0.107 |
| | 0.225 | NB | 137 | 25 | 1228 | 24 [9; 28] | 1338 [29; 2416] | 0.130 |
| | | Poisson | 130 | 25 | 1228 | 24 [10; 27] | 1082 [38; 1928] | 0.107 |
| **κ** | | | | | | | | |
| | 2 | NB | 125 | 19 | 219 | 17 [0; 29] | 443 [0; 1647] | 0.750 |
| | | Poisson | 120 | 18 | 178 | 16 [0; 28] | 341 [0; 1374] | 0.774 |
| | 3 (ref) | NB | 137 | 28 | 1347 | 27 [9; 31] | 1426 [25; 2459] | 0.130 |
| | | Poisson | 130 | 27 | 1102 | 26 [10; 30] | 1161 [32; 2012] | 0.107 |
| | 4 | NB | 149 | 31 | 2459 | 30 [26; 30] | 2314 [901; 3671] | 0.009 |
| | | Poisson | 140 | 30 | 2012 | 29 [25; 32] | 1889 [737; 3004] | 0.004 |

*Abbreviations*: FP: false positive rate; NB: negative binomial; 95%PI: 95% empirical predictive intervals derived from Monte Carlo simulations to estimate the probability of first detection; ref: reference value; $t_{md}$: median detection time; $t_E$: expected time to detection

[a] Time units correspond to the surveillance system's reporting period (daily or weekly in most jurisdictions) and need to be interpreted in the context of the monitored pathogen. For systems with daily cadence monitoring rapidly propagating pathogens (e.g., daily growth rates of 0.20), this represents 28-35 days to detection. For weekly reporting systems monitoring slower-spreading pathogens, this could represent weeks.

[b] False positive rate computed based on *s = 10,000* simulations

### Supplementary Figures

### Supplementary Figure S1

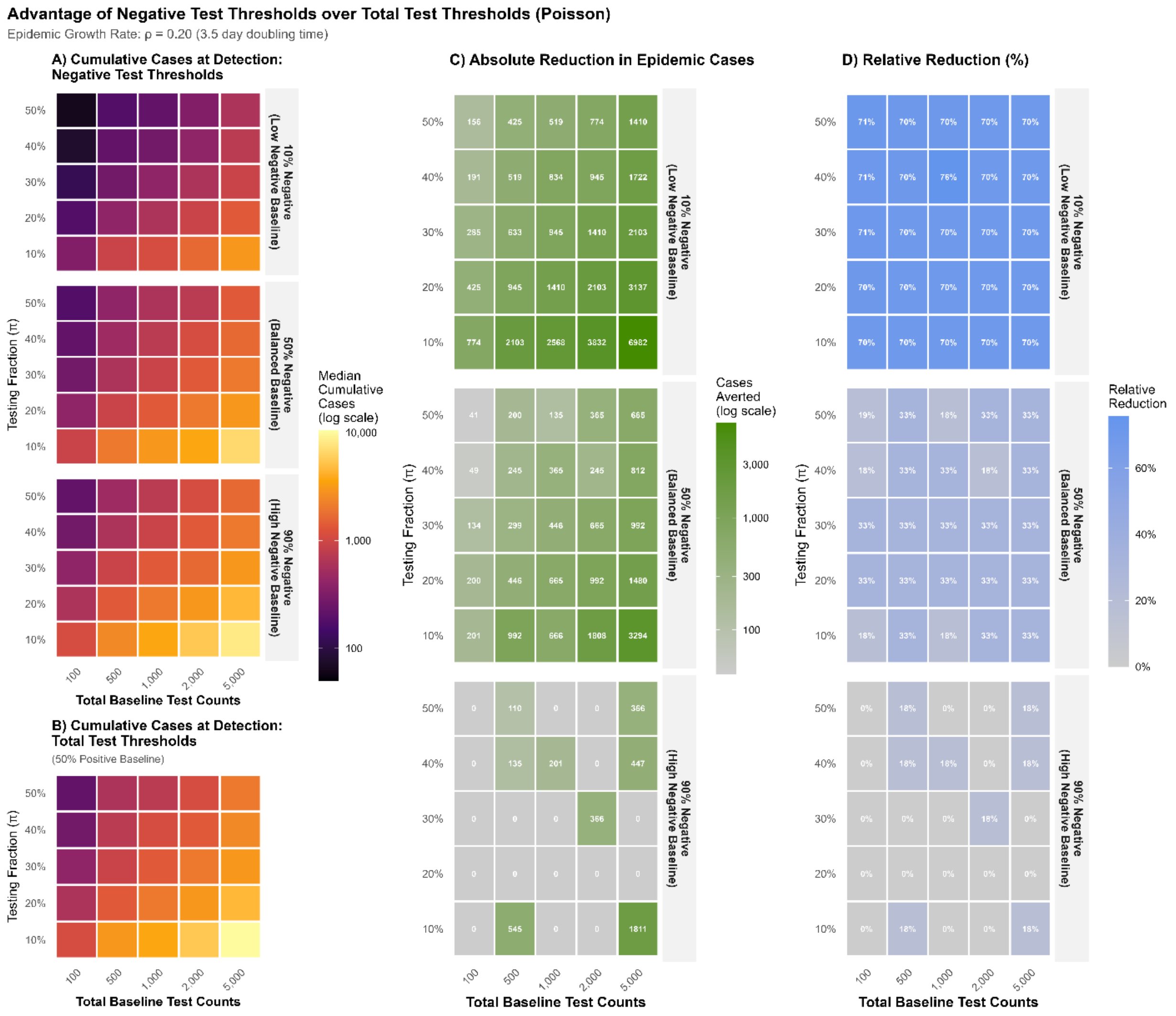


Heatmaps showing epidemic detection performance across surveillance parameters for two threshold strategies. **Panel A** (Negative Test Thresholds): Detection based on negative test counts for the routine condition. Color gradients indicate median cumulative epidemic cases at detection (log scale: dark purple = early detection with fewer cases; yellow = late detection with more cases). Rows represent increasing baseline proportions of negative tests, demonstrating performance across different baseline testing patterns. The x-axis shows total baseline test counts; the y-axis shows the testing fraction (π), representing the proportion of individuals with the epidemic condition who also receive the routine diagnostic test. **Panel B** (Total Test Thresholds): Detection based on total test counts. **Panel C** (Absolute Case Reduction): Absolute difference in median cumulative cases at detection between strategies. Darker colors (green) indicate larger benefit. **Panel D** (Relative Case Reduction): Percentage reduction in median cumulative cases at detection when using negative test thresholds compared to total test thresholds, calculated as (cases averted / cases at detection using total tests) × 100%. Positive percentages indicate proportional improvement from the negative test strategy. Rows show results stratified by baseline negative test proportions.

All panels display results for epidemic growth rate $\rho = 0.20$ (3.5-day doubling time). X-axes show total baseline test count; Y-axes show testing fraction ($\pi$).
See supplementary *Figure S2 – S5* for detailed heatmaps displaying estimates of median cumulative epidemic cases at detection.

### Supplementary Figure S2: Detailed Heatmaps for Negative Binomial Analytic Estimates

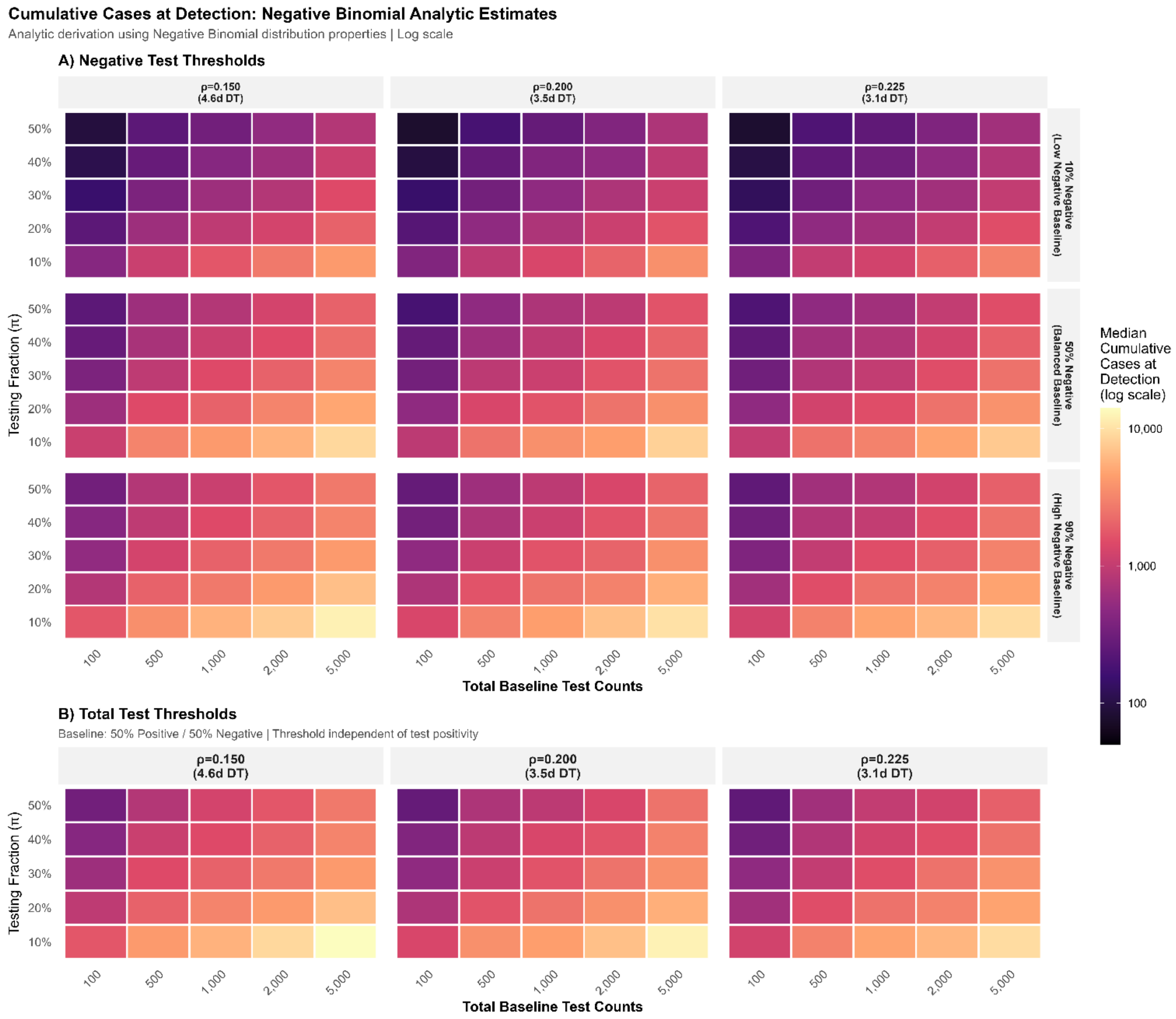


Heatmaps show epidemic detection performance across surveillance parameters for two threshold strategies. Color gradients indicate median cumulative epidemic cases at detection (log scale: dark purple = early detection with fewer cases; yellow = late detection with more cases). **Panel A**: Detections based on negative test thresholds. Rows show increasing proportions of negative tests; columns show increasing epidemic growth rates (ρ and doubling times (DT) in days (d)); X-axes show total baseline test counts; Y-axes show the testing fraction (π), representing the proportion of individuals with the epidemic condition who also receive the routine diagnostic test. **Panel B**: Detections based on total test thresholds. For thresholds based on total tests stratification by proportion of negative baseline test (righthand axis) was omitted because threshold exceedance depends solely on total test volume, so detection performance is invariant to the proportion of positive versus negative baseline tests.

### Supplementary Figure S3: Detailed Heatmaps for Poisson Analytic Estimates

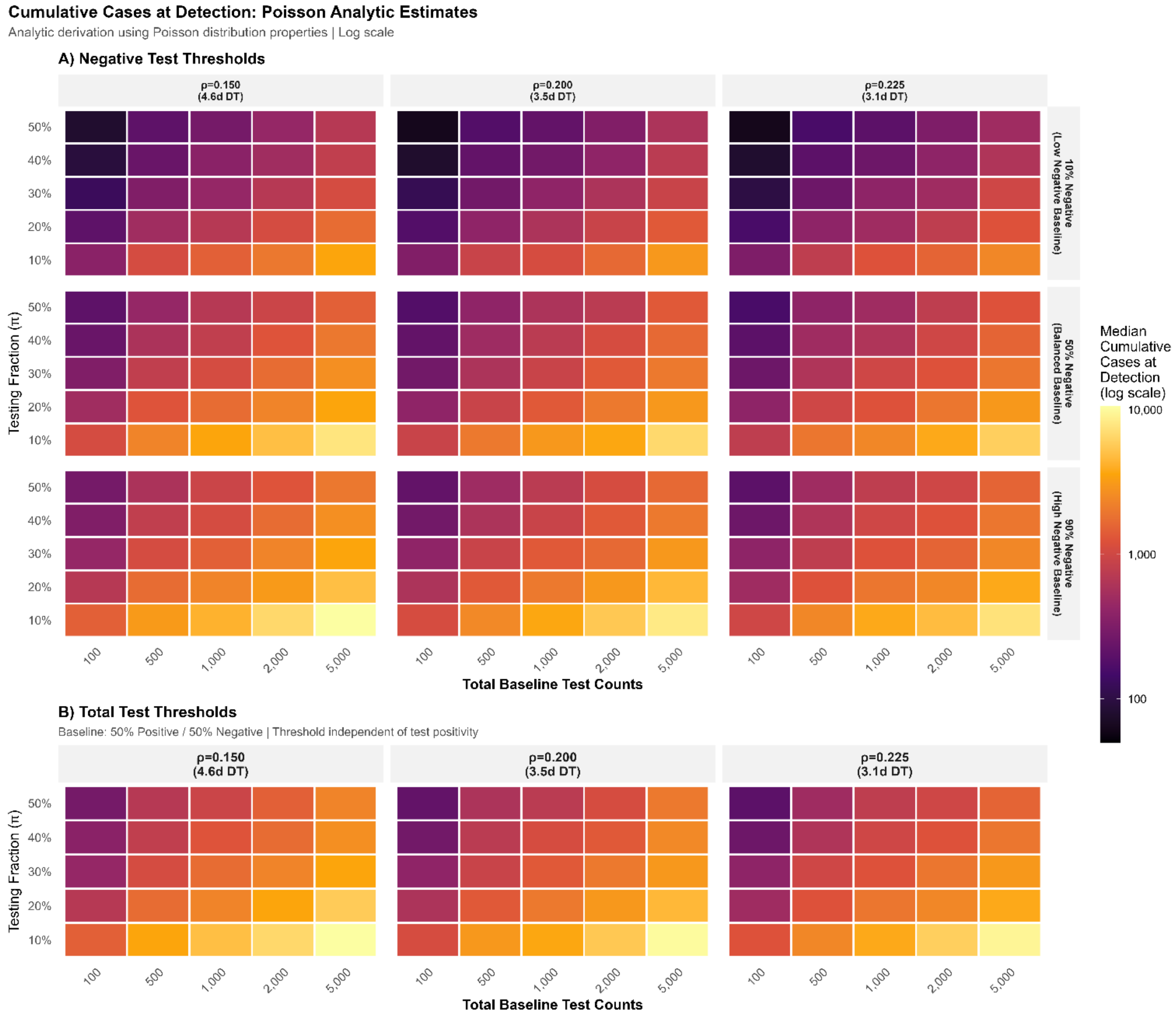


Heatmaps show epidemic detection performance across surveillance parameters for two threshold strategies. Color gradients indicate median cumulative epidemic cases at detection (log scale: dark purple = early detection with fewer cases; yellow = late detection with more cases). **Panel A**: Detections based on negative test thresholds. Rows show increasing proportions of negative tests; columns show increasing epidemic growth rates (ρ and doubling times (DT) in days (d)); X-axes show total baseline test counts; Y-axes show the testing fraction (π), representing the proportion of individuals with the epidemic condition who also receive the routine diagnostic test. **Panel B**: Detections based on total test thresholds. For thresholds based on total tests stratification by proportion of negative baseline test (righthand axis) was omitted because threshold exceedance depends solely on total test volume, so detection performance is invariant to the proportion of positive versus negative baseline tests.

### Supplementary Figure S4: Detailed Heatmaps for Negative Binomial Simulation Estimates

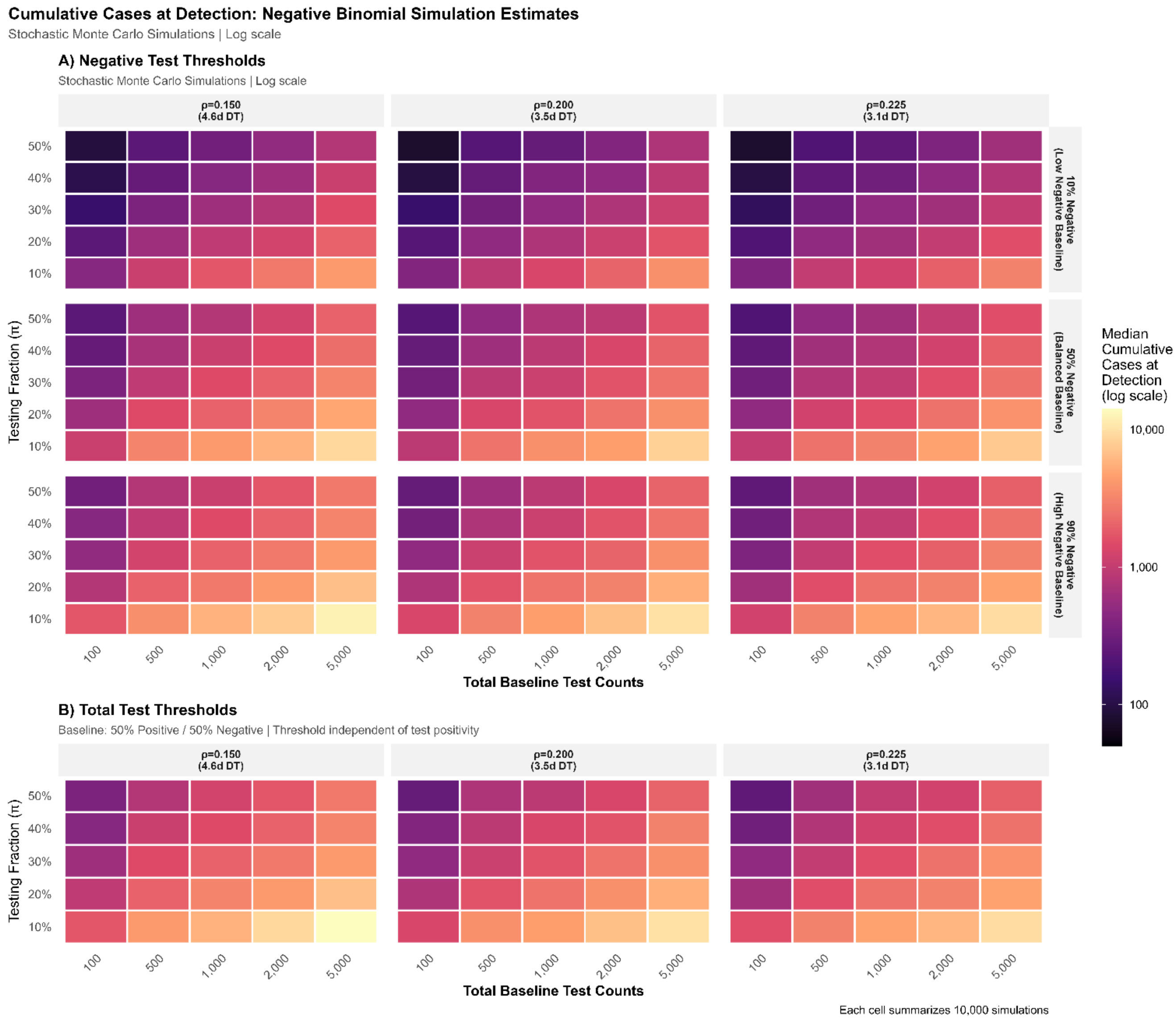


Heatmaps show epidemic detection performance across surveillance parameters for two threshold strategies. Color gradients indicate median cumulative epidemic cases at detection (log scale: dark purple = early detection with fewer cases; yellow = late detection with more cases). **Panel A**: Detections based on negative test thresholds. Rows show increasing proportions of negative tests; columns show increasing epidemic growth rates (ρ and doubling times (DT) in days (d)); X-axes show total baseline test counts; Y-axes show the testing fraction (π), representing the proportion of individuals with the epidemic condition who also receive the routine diagnostic test. **Panel B**: Detections based on total test thresholds. For thresholds based on total tests stratification by proportion of negative baseline test (righthand axis) was omitted because threshold exceedance depends solely on total test volume, so detection performance is invariant to the proportion of positive versus negative baseline tests.

### Supplementary Figure S5: Detailed Heatmaps for Poisson Simulation Estimates

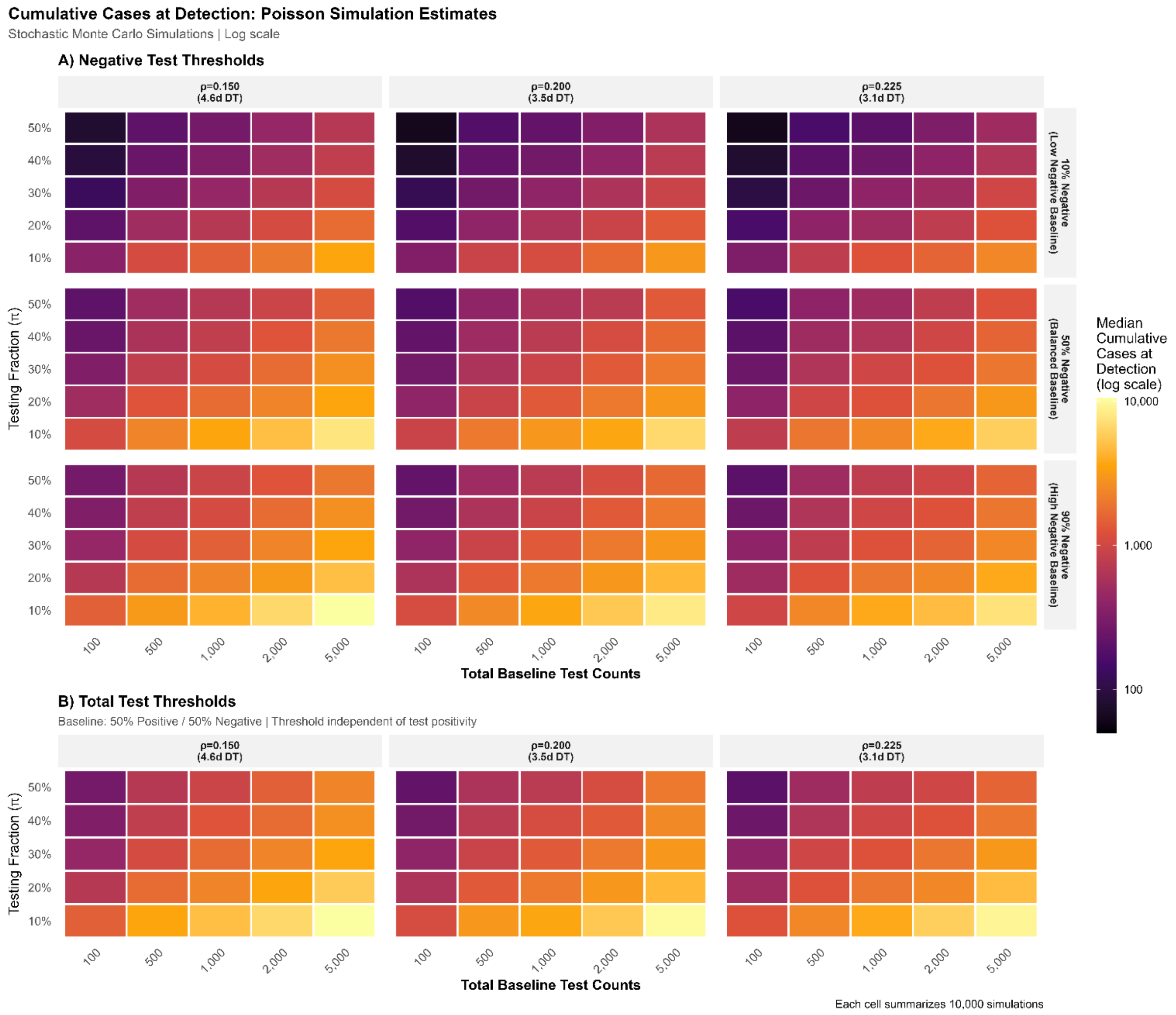


Heatmaps show epidemic detection performance across surveillance parameters for two threshold strategies. Color gradients indicate median cumulative epidemic cases at detection (log scale: dark purple = early detection with fewer cases; yellow = late detection with more cases). **Panel A**: Detections based on negative test thresholds. Rows show increasing proportions of negative tests; columns show increasing epidemic growth rates (ρ and doubling times (DT) in days (d)); X-axes show total baseline test counts; Y-axes show the testing fraction (π), representing the proportion of individuals with the epidemic condition who also receive the routine diagnostic test. **Panel B**: Detections based on total test thresholds. For thresholds based on total tests stratification by proportion of negative baseline test (righthand axis) was omitted because threshold exceedance depends solely on total test volume, so detection performance is invariant to the proportion of positive versus negative baseline tests.

### Supplementary Figure S6: Impact of Testing Fraction on Epidemic Size Estimates (pre vs. post-detection scenarios)

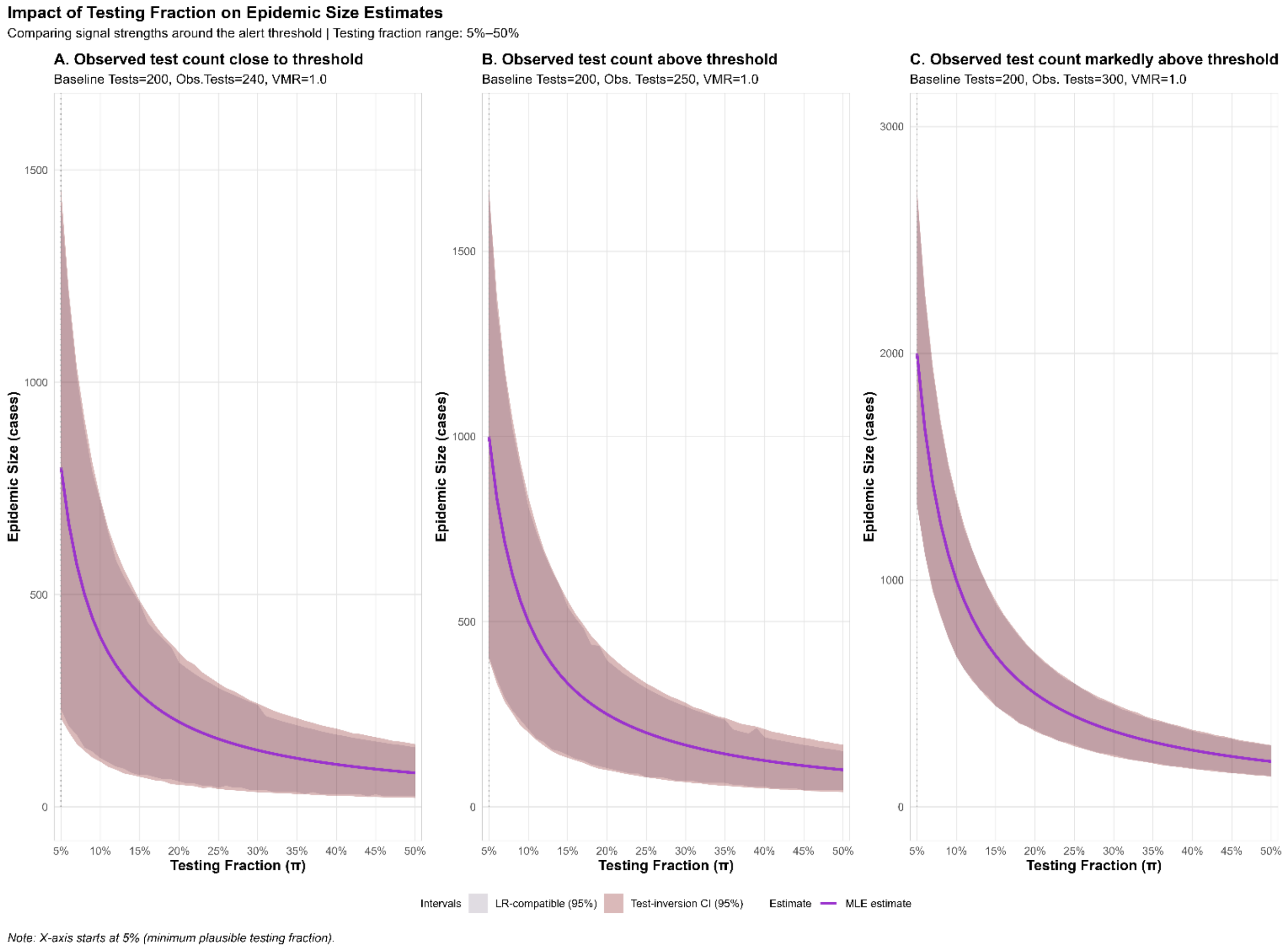


Comparison examining how epidemic size estimates change as observed test counts transition from below to above the detection threshold (at ~243 total tests). X-axis starts at 5% (minimum plausible π). **Solid lines**: Maximum likelihood estimate (MLE) of the underlying epidemic. **Shaded regions**: dark = 95% confidence interval, light = 95% likelihood ratio-compatible set. Panels compare: **(A)** Moderate excess signal approaching detection threshold (observed=240 vs. threshold=243); **(B)** Signal slightly above the detection threshold (observed=250 vs. threshold=243); **(C)** Signal substantially exceeding detection threshold (observed=300 vs. threshold=243). All scenarios use variance-to-mean ratio (VMR)=1.0 (Poisson baseline), κ=3 standard deviation threshold.

*Note*: X-axes are truncated at 5% (minimum plausible π) to improve readability (low testing fractions $\pi < 0.05$ heavily inflate the estimated epidemic size at detection resulting in very large y-axes). The detection approach is not practical for very low testing fractions ($\pi < 0.05$) given that only a very small fraction of the underlying emerging epidemic would be captured in the routine testing stream.

**Alt text for figures**

**Main text figures**

**Figure 1 alt text:** Four-panel heatmap comparing epidemic detection performance between negative test and total test threshold strategies under Negative Binomial assumptions accounting for overdispersion. Panel A shows median cumulative cases at detection using negative test thresholds across three baseline scenarios (10%, 50%, and 90% negative tests), with colors ranging from dark purple (early detection) to yellow (delayed detection with over 10,000 cumulative cases). Detection performance improves with higher testing fractions and lower baseline volumes. Panel B shows total test threshold performance, with consistently later detection than negative test thresholds (more yellow coloring). Panel C displays absolute cases averted, showing the greatest advantage (up to 8,000+ cases prevented) when baseline negative test proportion is low; this advantage largely disappears when negative tests dominate the baseline. Panel D shows relative case reduction of approximately 70% for low negative baseline scenarios, 33% for balanced baselines, and near zero for high negative baseline settings. Compared to Poisson results (Figure 1), Negative Binomial estimates show slightly later detection times overall, reflecting increased variance from overdispersion. All results shown for epidemic growth rate $\rho = 0.20$.

**Figure 2 alt text:** Four-panel line plot showing epidemic size estimates as a function of testing fractions ($\pi$) across different surveillance scenarios. All panels display a steep inverse relationship: maximum likelihood estimates (purple lines) decrease sharply as testing fractions increase from 5% to approximately 25%, then plateau at higher fractions. Shaded ribbons show 95% confidence intervals (darker) nested within 95% likelihood ratio-compatible sets (lighter); both uncertainty regions are extremely wide at low testing fractions (below 10%) and narrow substantially above 30%. Panel A (reference: baseline tests=200, observed tests =205) shows estimates ranging from approximately 700 cases at $\pi$=5% to under 100 cases at $\pi$=50%. Panel B (stronger signal: baseline tests =200, observed tests =240) shows larger estimates reaching approximately 1,400 cases at low $\pi$, with proportionally tighter (non-zero) confidence intervals. Panel C (low baseline tests =100) yields smaller absolute estimates but with large relative uncertainty. Panel D (high baseline=1,000) produces the largest absolute estimates (exceeding 2,000 cases at $\pi$=5%) but demonstrates more stable relative precision across the range of testing fractions. All scenarios assume Poisson baseline variability (VMR=1.0) and $\kappa$=3 detection threshold.

**Figure 3 alt text:** Four-panel plot showing how overdispersion affects epidemic size estimation across different surveillance scenarios. Each panel displays estimated epidemic size (y-axis) versus observed test count (x-axis), with five variance-to-mean ratio (VMR) levels represented by nested colored ribbons: purple ($\varphi$=1.0, Poisson), blue-purple ($\varphi$=1.5), teal ($\varphi$=2.0), green ($\varphi$=3.0), and yellow ($\varphi$=5.0). Solid lines show maximum likelihood estimates, which are identical across VMR levels (overlaid, with only $\varphi$=5.0 visible). Shaded ribbons show 95% confidence intervals (darker) and likelihood ratio-compatible sets (lighter), expanding substantially with increasing VMR—yellow ribbons are approximately twice as wide as purple. Colored dotted vertical lines mark detection thresholds for each VMR, shifting rightward with higher overdispersion to indicate delayed detection. A solid vertical line marks the baseline mean; gray shading indicates ±10% of baseline. For observed counts below or near baseline, confidence intervals are bound at zero. Panel A (baseline tests=200, $\pi$=10%) shows the reference scenario with estimates reaching 1,500+ cases at high observed counts. Panel B (baseline tests=200, $\pi$=50%) demonstrates how higher testing fractions ($\pi$) reduce estimated epidemic size and improve relative precision, with y-axis compressed to approximately 300 cases.

Panel C (low baseline tests=100) shows smaller absolute estimates with detection thresholds clustered more closely. Panel D (high baseline tests=1,000) illustrates the scale penalty, with estimates exceeding 6,000 cases and requiring proportionally stronger signals for detection. All results are based on 10,000-iteration simulations with $\kappa=3$ threshold.

#### Supplementary figures

**Figure S1 alt text:** Four-panel heatmap comparing epidemic detection performance between negative test and total test threshold strategies under Poisson assumptions. Panel A shows median cumulative cases at detection using negative test thresholds across three baseline scenarios (10%, 50%, and 90% negative tests), with colors ranging from dark purple (fewer than 100 cases, indicating early detection) to yellow (over 10,000 cases, indicating delayed detection). Best performance occurs at high testing fractions and low baseline test counts. Panel B shows the same metric for total test thresholds, displaying consistently later detection (more yellow coloring) than Panel A under equivalent conditions. Panel C displays absolute cases averted by using negative test thresholds, with dark green indicating thousands of cases prevented; the advantage is largest when baseline negative test proportion is low (10%) and diminishes to near zero when baseline negative tests are high (90%). Panel D shows relative case reduction, demonstrating consistent 70% improvement for low negative baseline scenarios, approximately 33% for balanced baselines, and minimal advantage when baseline negative tests predominate. All results shown for epidemic growth rate $\rho = 0.20$.

**Figure S2 alt text:** Two-panel heatmap array showing epidemic detection performance under Poisson analytic assumptions. Panel A displays negative test threshold results in a 3×3 grid: columns represent epidemic growth rates ($\rho=0.150$ with 4.6-day doubling time, $\rho=0.200$ with 3.5-day doubling time, $\rho=0.225$ with 3.1-day doubling time); rows represent baseline negative test proportions (10%, 50%, 90%). Each individual heatmap shows testing fractions (y-axis, 10%–50%) versus total baseline test counts (x-axis, 100–5,000), with color indicating median cumulative cases at detection on a log scale from dark purple (fewer than 100 cases, early detection) to yellow (over 10,000 cases, delayed detection). Detection performance improves (more purple) with higher growth rates, higher testing fractions, lower baseline volumes, and lower baseline negative proportions. Panel B shows total test threshold results in a single row of three heatmaps (one per growth rate), displaying consistently later detection (more yellow/orange coloring) compared to negative test thresholds in Panel A, particularly for low baseline negative proportion scenarios. The total test threshold results are independent of baseline positive/negative composition.

**Figure S3 alt text:** Two-panel heatmap array showing epidemic detection performance under Negative Binomial analytic assumptions accounting for overdispersion. Panel A displays negative test threshold results in a 3×3 grid: columns represent epidemic growth rates ($\rho=0.150$, 0.200, 0.225 corresponding to 4.6, 3.5, and 3.1-day doubling times); rows represent baseline negative test proportions (10%, 50%, 90%). Each heatmap shows testing fractions versus baseline test counts, with color indicating median cumulative cases at detection (log scale: dark purple = early detection, yellow = delayed detection). Compared to Poisson estimates (Figure S1), these heatmaps show slightly warmer (yellow/orange) tones overall, reflecting later detection times due to overdispersion in baseline test counts. The same parameter relationships hold: detection improves with higher growth rates, higher testing fractions, and lower baselines. Panel B shows total test threshold results across three growth rates, with detection consistently later than negative test thresholds. Pattern similarity to Figure S1 validates the analytic framework across distributional assumptions.

**Figure S4 alt text:** Two-panel heatmap array showing epidemic detection performance from Monte Carlo simulations under Poisson assumptions (10,000 iterations per parameter combination). Panel A displays negative test threshold results in a 3×3 grid organized by epidemic growth rate (columns: ρ=0.150, 0.200, 0.225) and baseline negative test proportion (rows: 10%, 50%, 90%). Color gradients indicate median cumulative cases at detection (log scale: dark purple = fewer cases at detection, yellow = more cases). Detection performance follows consistent patterns: earlier detection (more purple) occurs with faster epidemic growth, higher testing fractions, lower baseline volumes, and lower baseline negative proportions. Panel B shows total test threshold results in a single row, displaying later detection compared to negative test thresholds. The simulation results are visually nearly identical to Poisson analytic estimates (Figure S1), validating agreement between analytical and simulation methodologies. Each cell summarizes 10,000 stochastic simulations.

**Figure S5 alt text:** Two-panel heatmap array showing epidemic detection performance from Monte Carlo simulations under Negative Binomial assumptions accounting for overdispersion (10,000 iterations per parameter combination). Panel A displays negative test threshold results in a 3×3 grid: columns show epidemic growth rates (ρ=0.150, 0.200, 0.225); rows show baseline negative test proportions (10%, 50%, 90%). Color indicates median cumulative cases at detection on a log scale from dark purple (early detection) to yellow (delayed detection). Compared to Poisson simulations (Figure S3), these heatmaps show slightly later detection times (warmer colors) due to increased baseline variability. Panel B shows total test threshold results, with consistently later detection than negative test thresholds in Panel A. The close visual correspondence between simulation results (Figures S3 & S4) and analytic estimates (Figures S1 & S2) across both Poisson and Negative Binomial assumptions demonstrates strong methodological agreement and validates the analytical framework.

**Figure S6 alt text:** Three-panel line plot comparing epidemic size estimates as observed test counts transition from below to above the detection threshold (approximately 243 tests). All panels show testing fractions (π) on the x-axis (5%–50%) and estimated epidemic size on the y-axis, with maximum likelihood estimates as purple lines and shaded uncertainty regions for 95% confidence intervals (darker) and likelihood ratio-compatible sets (lighter). Panel A (observed tests=240, just below threshold) shows estimates declining from approximately 800 cases at π=5% to under 200 cases at π=50%, with wide confidence intervals especially at low testing fractions. Panel B (observed tests=250, slightly above threshold) displays a similar curve shape with modestly larger estimates and comparably wide uncertainty. Panel C (observed=300, substantially above threshold) shows the largest epidemic size estimates, reaching approximately 2,000 cases at low testing fractions, but demonstrates markedly improved precision—confidence intervals are visibly narrower relative to the point estimates across all testing fractions, including at low π values where Panels A and B show substantial uncertainty. This illustrates that crossing the detection threshold substantially improves epidemic size estimation precision. All scenarios use baseline=200 tests, VMR=1.0 (Poisson), and κ=3 threshold.

**Used R packages**

*R version:*

platform x86_64-w64-mingw32

arch x86_64

os mingw32

crt ucrt

system x86_64, mingw32

status

major 4

minor 5.1

year 2025

month 06

day 13

svn rev 88306

language R

version.string R version 4.5.1 (2025-06-13 ucrt)

nickname Great Square Root

*Packages*:

Garnier, Simon, Ross, Noam, Rudis, Robert, Camargo, Pedro A, Sciaini, Marco, Scherer, Cédric (2024). _viridis(Lite) - Colorblind-Friendly Color Maps for R_. doi:10.5281/zenodo.4679423 <https://doi.org/10.5281/zenodo.4679423>, viridis package version 0.6.5, <https://sjmgarnier.github.io/viridis/>.

Makowski D, Lüdecke D, Patil I, Thériault R, Ben-Shachar M, Wiernik B (2023). "Automated Results Reporting as a Practical Tool to Improve Reproducibility and Methodological Best Practices Adoption." _CRAN_. doi:10.32614/CRAN.package.report <https://doi.org/10.32614/CRAN.package.report>, <https://easystats.github.io/report/>.

Pedersen T (2025). _patchwork: The Composer of Plots_. doi:10.32614/CRAN.package.patchwork <https://doi.org/10.32614/CRAN.package.patchwork>, R package version 1.3.1, <https://CRAN.R-project.org/package=patchwork>.

R Core Team (2025). _R: A Language and Environment for Statistical Computing_. R Foundation for Statistical Computing, Vienna, Austria. <https://www.R-project.org/>.

Urbanek S, Horner J (2025). _Cairo: R Graphics Device using Cairo Graphics Library for Creating High-Quality Bitmap (PNG, JPEG, TIFF), Vector (PDF, SVG, PostScript) and Display (X11 and Win32) Output_. doi:10.32614/CRAN.package.Cairo <https://doi.org/10.32614/CRAN.package.Cairo>, R package version 1.7-0, <https://CRAN.R-project.org/package=Cairo>.

Wickham H (2016). _ggplot2: Elegant Graphics for Data Analysis_. Springer-Verlag New York. ISBN 978-3-319-24277-4, <https://ggplot2.tidyverse.org>.

Wickham H, François R, Henry L, Müller K, Vaughan D (2023). _dplyr: A Grammar of Data Manipulation_. doi:10.32614/CRAN.package.dplyr <https://doi.org/10.32614/CRAN.package.dplyr>, R package version 1.1.4, <https://CRAN.R-project.org/package=dplyr>.

Wickham H, Pedersen T, Seidel D (2025). _scales: Scale Functions for Visualization_. doi:10.32614/CRAN.package.scales <https://doi.org/10.32614/CRAN.package.scales>, R package version 1.4.0, <https://CRAN.R-project.org/package=scales>.

Wickham H, Vaughan D, Girlich M (2024). _tidyr: Tidy Messy Data_. doi:10.32614/CRAN.package.tidyr <https://doi.org/10.32614/CRAN.package.tidyr>, R package version 1.3.1, <https://CRAN.R-project.org/package=tidyr>.
